# Comparing the genetic and environmental architecture of blood count, blood biochemistry and urine biochemistry biological ages with machine learning

**DOI:** 10.1101/2021.07.05.21260032

**Authors:** Alan Le Goallec, Samuel Diai, Théo Vincent, Chirag J. Patel

## Abstract

While a large number of biological age predictors have been built from blood samples, a blood count-based biological age predictor is lacking, and the genetic and environmental factors associated with blood-measured accelerated aging remain elusive. In the following, we leveraged 31 blood count biomarkers measured from 489,079 blood samples, 28 blood biochemistry biomarkers measured from 245,147 blood samples, and four urine biochemistry biomarkers measured from 158,381 samples to build three distinct biological age predictors by training machine learning models to predict age. Blood biochemistry significantly outperformed blood count and urine biochemistry in terms of age prediction (RMSE: 5.92+-0.02 vs. 7.60+-0.02 years and 7.72+-0.04 years). We performed genome wide association studies [GWASs], and found accelerated blood biochemistry, blood count and urine biochemistry aging to be respectively 26.2+-0.3%, 18.1+-0.2% and 10.5±0.5% GWAS-heritable. We identified 1,081 single nucleotide polymorphisms [SNPs] associated with accelerated blood biochemistry aging, 2,636 SNPs associated with accelerated blood cells aging and 24 SNPs associated with accelerated urine biochemistry aging. Similarly, we identified biomarkers, clinical phenotypes, diseases, environmental and socioeconomic factors associated with accelerated blood biochemistry, blood cells and urine biochemistry aging.

## Introduction

With the world population aging ^1^, the prevalence of age-related diseases such as cardiovascular disease, cancer, osteoarthritis, type 2 diabetes, osteoporosis, Parkinson’s disease and Alzheimer’s disease is projected to increase ^2^, limiting the gains in life expectancy ^3^. To better understand aging, biological age predictors have been developed by training machine learning models to predict age (also referred to as “chronological age”). After the models have been trained, the prediction they output on unseen samples can be interpreted as the “biological age” of the participant. Participants whose biological age is higher than their chronological age are called accelerated agers. Biological age predictors have been developed from diverse data modalities ^4^, including blood ^5–11^ and urine ^12^ biochemistry biomarkers.

In the following, we leveraged laboratory biomarkers collected from 37-82 year-old UK Biobank [UKB] ^13^ participants to build a blood cells age predictor, a blood biochemistry age predictor and a urine biochemistry age predictor. Specifically, we used 31 blood count biomarkers measured from 489,079 blood samples, 28 blood biochemistry biomarkers measured from 245,147 blood samples, and four urine biochemistry biomarkers measured from 158,381 urine samples. UKB’s laboratory biomarkers (blood count, blood biochemistry and urine biochemistry) have previously been analyzed to predict survival ^14,15^, but we are, to our knowledge, the first to leverage them to predict chronological age. We defined accelerated aging for each of these three biological age dimensions as the difference between accelerated aging and chronological age. For example, a 50 year-old participant for which the blood biochemistry biomarkers-based model predicted an age of 60 years has a blood biochemistry age of 60 years and is a ten years accelerated ager in terms of blood biochemistry. We then performed three genome wide association studies [GWASs] to identify the heritability and single nucleotide polymorphisms [SNPs] associated with accelerated aging in each dimension. Similarly, we performed three X-Wide Association Studies [XWASs] to identify biomarkers, clinical phenotypes, diseases, environmental and socioeconomic variables associated with these three accelerated aging phenotypes. Finally, we quantified the correlation of these accelerated aging phenotypes at the phenotypic, genetic and environmental levels. (Figure 1)

**Figure 1:**
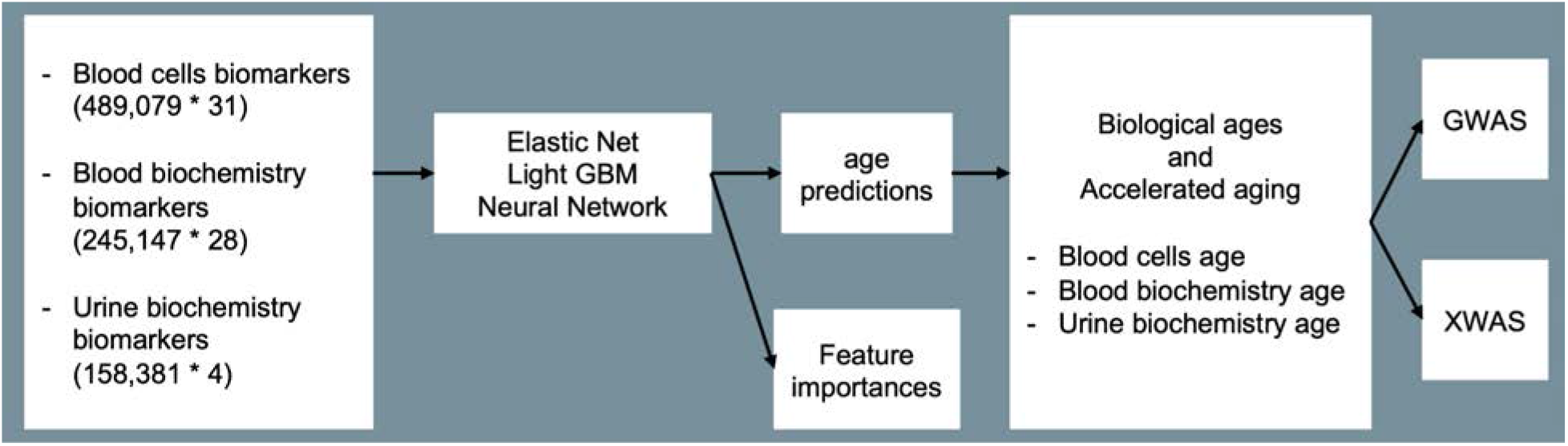
Overview of the datasets and analytic pipeline.

## Results

### UKB blood biochemistry biomarkers outperformed urine biochemistry and blood cell biomarkers as age predictors

We predicted chronological age from blood cell, blood biochemistry and urine biochemistry biomarkers using an ensemble of an elastic net, a gradient boosted machine [GBM] and a shallow, fully connected neural network. Blood biochemistry predicted chronological age with a R-Squared [R^2^] of 48.6+-0.4% and a root mean squared error of 5.92+-0.02 years, significantly outperforming blood cells (R^2^=12.5+-0.2%; RMSE=7.60+-0.02 years) and urine biochemistry (R^2^=10.4+-0.3%; RMSE=7.72+-0.04 years) (Figure 2 and Table S1). For the three datasets, the GBM and the neural network [NN] performed similarly and both outperformed the elastic net. For example, for blood cells, the GBM explained twice more variance in chronological age than the elastic net (R^2^=12.2+-0.2% vs. 5.8+-0.4%).

**Figure 2:**
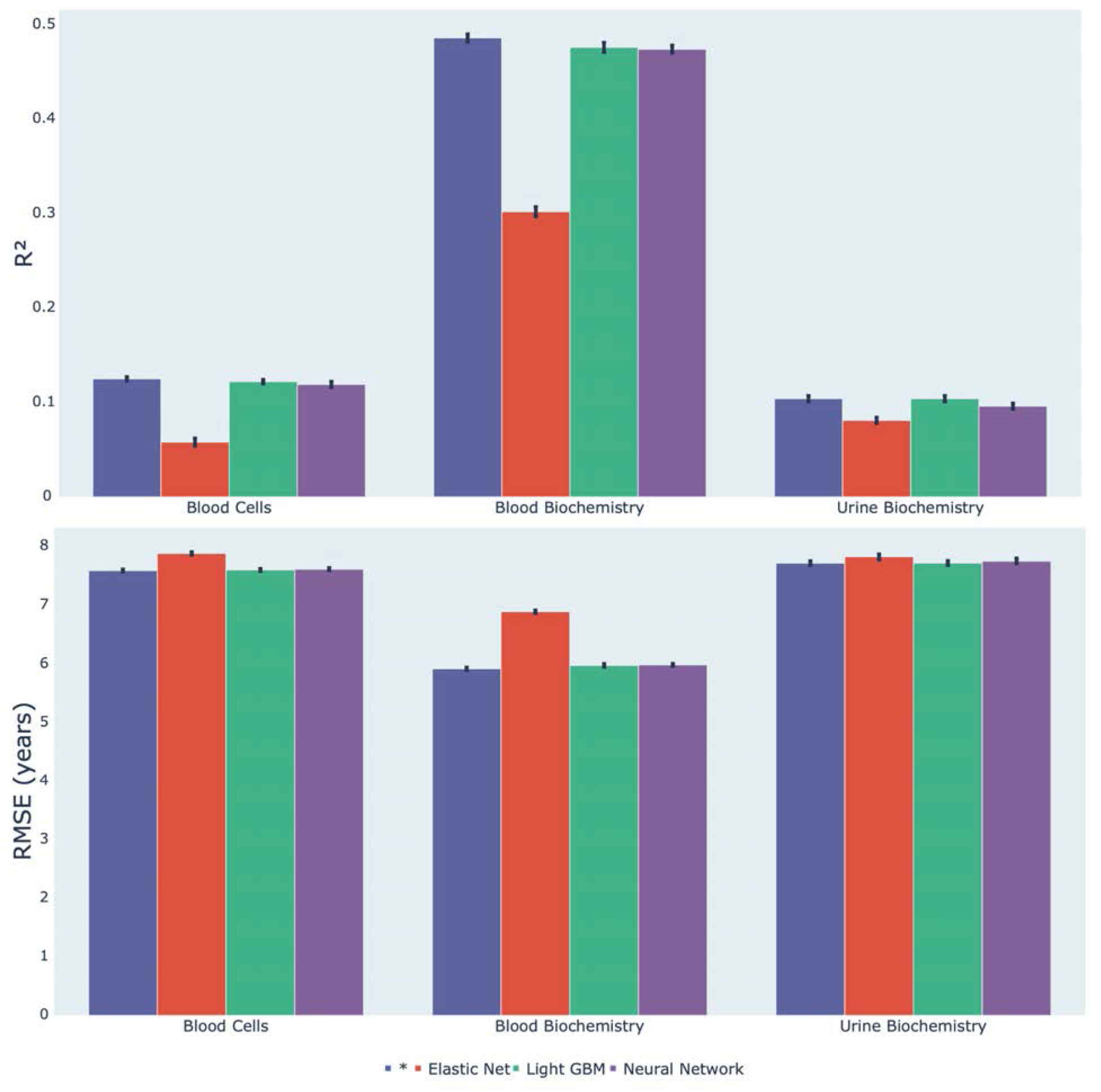
Chronological age prediction performance (R^2^ and RMSE) * represent ensemble models

### Identification of the laboratory biomarkers driving age prediction

The complete feature importances are found in Table S2, Table S3 and Table S4. Below, we summarize the most important findings for each aging dimension.

#### Blood biochemistry

We best predicted accelerated blood biochemistry aging using a GBM (R^2^=47.6±0.5%). The model was trained on 28 biomarkers, along with sex and ethnicity. Specifically, the most important biomarkers included (1) sex hormone binding globulin (SHBG), (2) testosterone, (3) glycated hemoglobin (HbA1c), (4) apolipoprotein B, (5) insulin-like growth factor 1 [IGF-1], (6) cystatin C, (7) direct low-density lipoprotein [LDL], (8) urea, (9) creatinine and (10) alanine aminotransferase. The elastic net (R^2^=30.2±0.5%) assigned a positive regression coefficient to sex hormone binding globulin (SHBG), glycated hemoglobin (HbA1c), apolipoprotein B, insulin-like growth factor 1 [IGF-1], cystatin C and urea, and a negative regression coefficient to testosterone, insulin-like growth factor 1 [IGF-1], direct low-density lipoprotein DL, creatinine and alanine aminotransferase.

#### Blood cells

We best predicted accelerated blood cells aging with a GBM (R^2^=12.2±0.2%) trained on 31 features, incorporating with sex and ethnicity in the predictor. The most predictive scalar features included (1) red blood cell distribution width, (2) red blood cell count, (3) mean corpuscular volume, (4) mean sphered volume, (5) platelet crit, (6) lymphocyte percentage, (7) mean reticulocyte volume, (8) neutrophil count, (9) hemoglobin concentration and (10) mean corpuscular hemoglobin concentration. The elastic net (R^2^=5.8±0.4%) assigned a positive regression coefficient to red blood cell distribution width, mean corpuscular volume, mean sphered cell volume, mean reticulocyte volume, neutrophil count and mean corpuscular hemoglobin concentration and a negative regression coefficient to red blood cell count, platelet crit, lymphocyte percentage and hemoglobin concentration.

#### Urine biochemistry

We best predicted accelerated urine biochemistry aging using a GBM (R^2^=10.4±0.3%). The model was trained on four biomarkers, along with sex and ethnicity. The most predictive scalar features included (1) creatinine, (2) potassium, (3) sodium, (4) microalbumin, (5) sex, (6) white ethnicity, (7) White - other, (8) ethnicity: prefer not to answer, (9) British and (10) Carribean. The elastic net (R^2^=8.1±0.3%) assigned a positive regression coefficient to potassium, microalbumin, sex, white ethnicity, ethnicity: prefer not to answer, and British and a negative regression coefficient to creatinine, sodium, and White - other ethnicity.

### Genetic factors and heritability of accelerated aging

We performed three genome wide association studies [GWASs] to estimate the GWAS-based heritability of blood biochemistry (h_g^2^=26.2±0.3%), blood cells (h_g^2^=18.1±0.2%), and urine biochemistry (h_g^2^=10.5±0.5%) accelerated aging, and to identify SNPs associated with these phenotypes (Figure 3 and Table 1).

**Figure 3:**
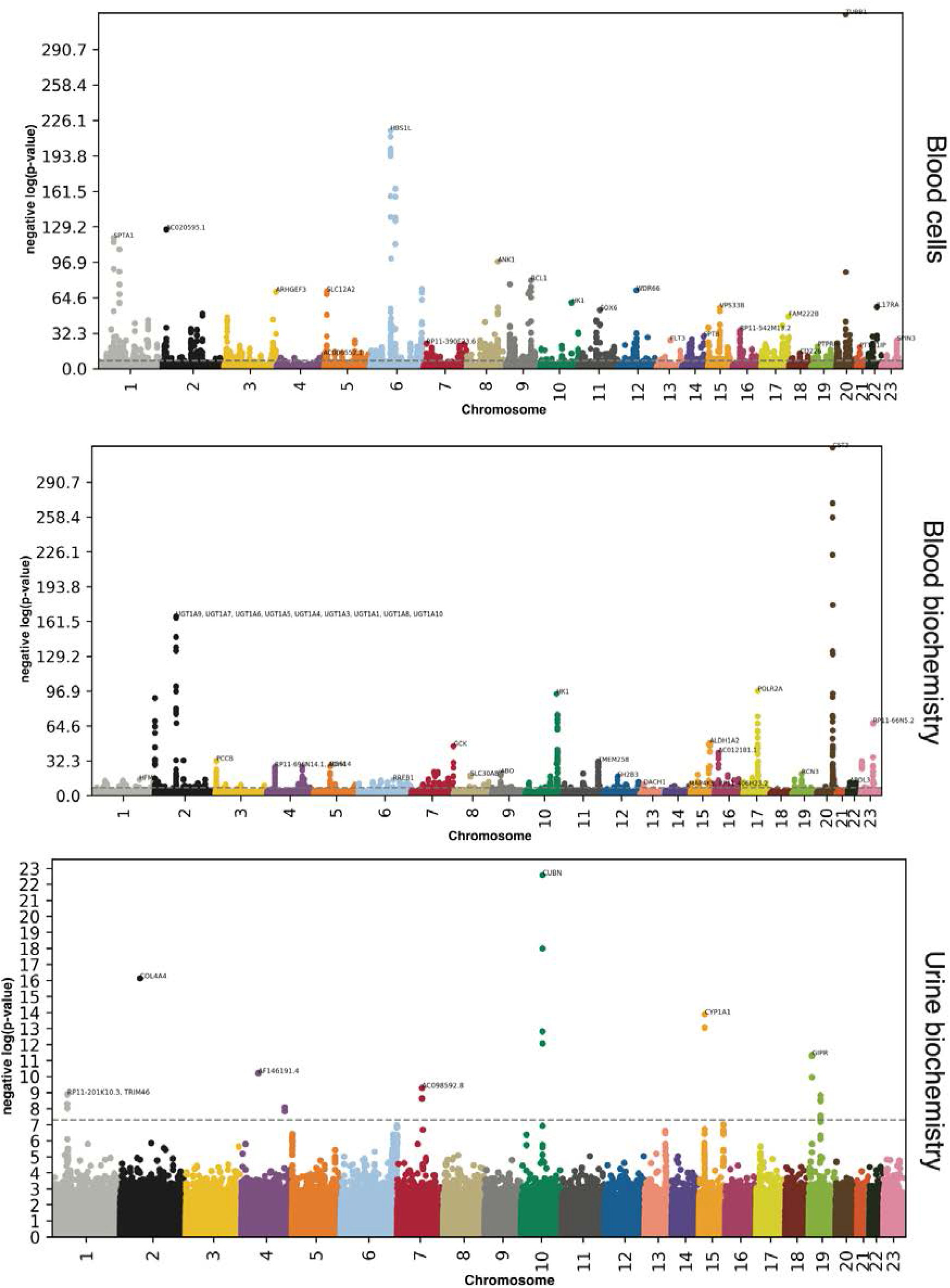
GWAS results - SNPs associated with accelerated aging for each dimension. −log10(p-value) vs. chromosomal position of locus. Dotted line denotes 5×10^−8^.

**Table 1:**
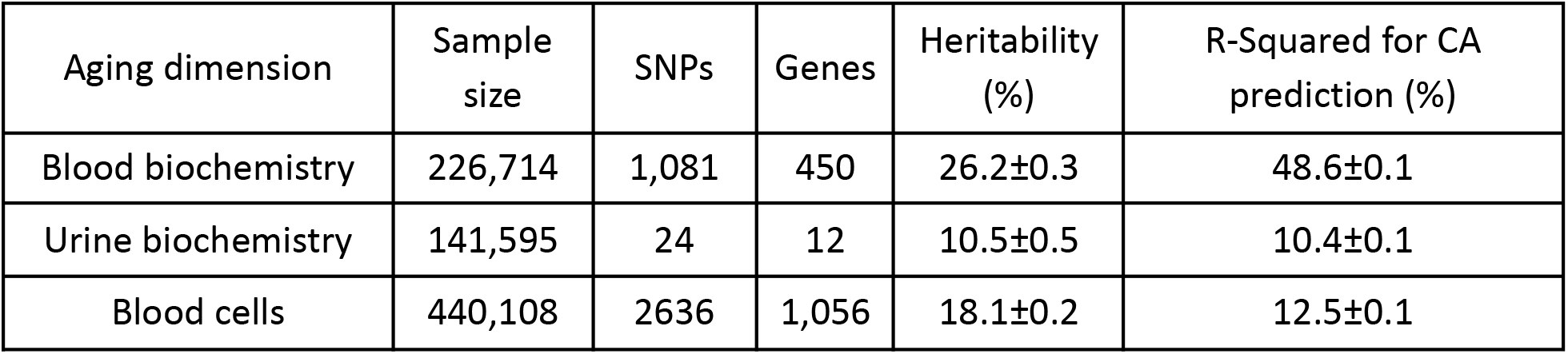
GWASs summary - Heritability, number of SNPs and genes associated with accelerated aging in each aging dimension.

#### Blood biochemistry

Accelerated blood biochemistry aging is 26.2±0.3% heritable and 1,081 SNPs in 450 genes are significantly associated with this phenotype. The ten highest GWAS peaks included loci in: (1) *CST3* (cystatin C); (2) *UDP* glucuronosyltransferase encoding genes (involved in drug metabolism); (3) *POLR2A* (RNA polymerase II, involved in mRNA synthesis); (4) HK1 (Hexokinase 1, involved in glucose metabolism); (5) *G6PC2* (Glucose-6-Phosphatase Catalytic Subunit 2, involved in glucose metabolism); (6) *RP11-66N5.2* (a pseudogene); (7) *ALDH1A2* (Aldehyde Dehydrogenase 1 Family Member A2, involved in the synthesis of retinoic acid); (8) GCK (Glucokinase, involved in glucose metabolism); (9) *AC012181.1* (a microRNA). AC012181.1 is in linkage disequilibrium with *HERPUD1* (Homocysteine Inducible ER Protein With Ubiquitin Like Domain 1, involved in ubiquitinated protein degradation) and *CETP* (Cholesteryl Ester Transfer Protein, involved in cholesterol metabolism); and (10) *TMEM258* (Transmembrane Protein 258, involved in protein N-glycosylation).

#### Blood cells

Accelerated blood cell aging is 18.1±0.2% heritable and 2,636 SNPs in 1,056 genes are significantly associated with this phenotype. The ten highest peaks highlighted by the GWAS are (1) *TUBB1* (Tubulin Beta 1 Class VI, involved in microtubule formation, expressed in thrombocytes and megakaryocytes and linked to macrothrombocytopenia); (2) *HBS1L* (*HBS1* Like Translational GTPase, involved in erythrocyte volume, hemoglobin content, erythrocyte count, platelet count and monocyte count, and linked to sickle cell disease and beta-thalassemia); (3) *AC020595.1* (a long intergenic non-coding RNA); (4) *SPTA1* (Spectrin Alpha, Erythrocytic 1, a component of red blood cell plasma membrane and linked to erythrocyte diseases such as elliptocytosis 2, pyropoikilocytosis, and type 3 spherocytosis); (5) *ANK1* (Ankyrin 1, Erythrocytic, a protein attaching membrane proteins to cytoskeletal elements, linked to spherocytosis - a red blood cell disease); (6) *RCL1* (RNA Terminal Phosphate Cyclase Like 1, involved in RNA processing and linked to breast lipoma); (7) *RP11-202G18.1* (a long intergenic non-coding RNA); (8) *WDR66* (Cilia And Flagella Associated Protein 251, involved in mean platelet volume and spermatozoa motility, and linked to spermatogenic failure and non-syndromic male infertility due to sperm motility disorder); (9) *SLC12A2* (Solute Carrier Family 12 Member 2, involved in sodium and chloride transport); and (10) *ARHGEF3* (Rho Guanine Nucleotide Exchange Factor 3, a guanine nucleotide exchange factor for RhoA and RhoB GTPases).

#### Urine biochemistry

Accelerated urine biochemistry aging is 10.5±0.5% heritable and 24 SNPs in 12 genes are significantly associated with this phenotype. Eight GWAS peaks can be identified, from most significantly associated to least significantly associated: (1) *CUBN* (cubilin, an endocytic receptor involved in vitamin B12, lipoprotein and iron metabolism) ^16^; (2) *COL4A4* (Collagen Type IV Alpha 4 Chain, a component of collagen that is only found in basement membranes) ^16^; (3) *CYP1A1* and *CYP1A2* (Cytochrome P450 Family 1 Subfamily A Members 1 and 2, involved in drug metabolism) ^16^; (4) *GIPR* (Gastric Inhibitory Polypeptide Receptor, stimulates insulin release when blood glucose is high) ^16^. *GIPR* is in linkage disequilibrium with FUT2 (Fucosyltransferase 2, a Golgi stack protein involved in A and B antigen synthesis ^16^); (5) AF146191.4 (5) AF146191.4 (long intergenic non-coding RNA with unknown function); (6) AC098592.8 (pseudogene with unknown function); (7) *TRIM46* (Tripartite Motif Containing 46, involved in microtubules fasciculation in the axon) ^16^. *TRIM46* is in linkage disequilibrium with RP11-201K10.3 (a protein coding gene with unknown function), *MUC1* (Mucin 1, Cell Surface Associated, involved in mucous secretion), *THBS3* (Thrombospondin 3, a glycoprotein involved in cell-to-cell and cell-to-matrix interactions) ^16^ and *MTX1* (Metaxin 1, involved in mitochondrial protein import); and (8) *FGF5* (Fibroblast Growth Factor 5, involved in cell mitogenesis and survival).

### Biomarkers, clinical phenotypes, diseases, environmental and socioeconomic variables associated with accelerated aging

We use “X” to refer to all nongenetic variables measured in the UK Biobank (biomarkers, clinical phenotypes, diseases, family history, environmental and socioeconomic variables). We performed an X-Wide Association Study [XWAS] to identify which of the 4,372 biomarkers classified in 21 subcategories (Table S7), 187 clinical phenotypes classified in 11 subcategories (Table S10), 2,073 diseases classified in 26 subcategories (Table S13), 92 family history variables (Table S16), 265 environmental variables classified in nine categories (Table S19), and 91 socioeconomic variables classified in five categories (Table S22) are associated (p-value threshold of 0.05 and Bonferroni correction) with accelerated aging in the different dimensions. We summarize our findings for general accelerated blood cells, blood biochemistry,and urine biochemistry aging in Table S8, Table S9, Table S11, Table S12, Table S14, Table S15, Table S20, Table S21, Table S23, Table S24, where we list the three variables of each X-subcategory (e.g spirometry biomarkers) most associated with each accelerated aging dimension. The full results can be exhaustively explored at https://www.multidimensionality-of-aging.net/xwas/univariate_associations.

Below, we provide an overview of the X-subcategories with a high proportion of variables associated with accelerated blood biochemistry aging.

#### Biomarkers associated with accelerated blood biochemistry aging

The three biomarker categories most associated with accelerated blood biochemistry aging, (blood biochemistry biomarkers themselves excepted) are urine biochemistry, blood count and arterial stiffness (Table S8). Specifically, 100.0% of urine biomarkers are associated with accelerated blood biochemistry aging, with the three largest associations being with creatinine (correlation=.083), sodium (correlation=.076), and potassium (correlation=.070). 61.3% of blood count biomarkers are associated with accelerated blood biochemistry aging, with the three largest associations being with eosinophil count (correlation=.076), white blood cell count (correlation=.075), and red blood cell distribution width (correlation=.069). 50.0% of pulse wave analysis-derived arterial stiffness biomarkers are associated with accelerated blood biochemistry aging, with the three largest associations being with position of the pulse wave peak (correlation=.045), position of the shoulder on the pulse waveform (correlation=.043), and absence of notch position in the pulse waveform (correlation=.037).

Conversely, the three biomarker categories most associated with decelerated blood biochemistry aging are symbol digit substitution (a cognitive test), body impedance and hand grip strength (Table S9). Specifically, 100.0% of symbol digit cognitive tests are associated with decelerated blood biochemistry aging, with the two associations being with number of digits matches made correctly (correlation=.045) and number of symbol digit matches attempted (correlation=.040). 100.0% of impedance biomarkers are associated with decelerated blood biochemistry aging, with the three largest associations being with left leg impedance (correlation=.072), right leg impedance (correlation=.069), and whole body impedance (correlation=.044). 100.0% of hand grip strength biomarkers are associated with decelerated blood biochemistry aging, with the two associations being with right hand grip strength (correlation=.077) and left hand grip strength (correlation=.076).

#### Clinical phenotypes associated with accelerated blood biochemistry aging

The three clinical phenotype categories most associated with accelerated blood biochemistry aging are chest pain, breathing and claudication (Table S11). Specifically, 100.0% of chest pain phenotypes are associated with accelerated blood biochemistry aging, with the three largest associations being with chest pain due to walking ceases when standing still (correlation=.052), chest pain or discomfort walking normally (correlation=.052), and chest pain or discomfort (correlation=.041). 100.0% of breathing phenotypes are associated with accelerated blood biochemistry aging, with the two associations being with shortness of breath walking on level ground (correlation=.092) and wheeze or whistling in the chest in the last year (correlation=.072). 76.9% of claudication phenotypes are associated with accelerated blood biochemistry aging, with the three largest associations being with leg pain when walking uphill or hurrying (correlation=.078), leg pain on walking (correlation=.075), and leg pain when standing still or sitting (correlation=.068).

Conversely, the three clinical phenotype categories most associated with decelerated blood biochemistry aging are cancer screening (ever had prostate specific antigen test: correlation=.030; ever had bowel cancer screening: correlation=.013), sexual factors (age first had sexual intercourse: correlation=.035) and oral health (no oral/dental problems: correlation=.035). (Table S12)

#### Diseases associated with accelerated blood biochemistry aging

The three disease categories most associated with accelerated blood biochemistry aging are cardiovascular disease, gastrointestinal tract disease, and a category involving diverse disorders (Table S14). Specifically, 57.1% of cardiovascular diseases are associated with accelerated blood biochemistry aging, with the three largest associations being with hypertension (correlation=.100), chronic ischaemic heart disease (correlation=.068), and angina pectoris (correlation=.055). 50.0% of gastrointestinal tract diseases are associated with accelerated blood biochemistry aging, with the three largest associations being with gastritis and duodenitis (correlation=.035), noninfective gastro-enteritis and colitis (correlation=.031), and diaphragmatic hernia (correlation=.028). 47.7% of the diseases in the diverse category are associated with accelerated blood biochemistry aging, with the three largest associations being with pain in the throat and chest (correlation=.045), abnormalities of breathing (correlation=.033), and nausea and vomiting (correlation=.027). Conversely, the three disease categories most associated with decelerated blood biochemistry aging are related to pregnancy and delivery (Table S15).

#### Environmental variables associated with accelerated blood biochemistry aging

The three environmental variable categories most associated with accelerated blood biochemistry aging are smoking, sleep and sun exposure (Table S20). Specifically, 50.0% of smoking variables are associated with accelerated blood biochemistry aging, with the three largest associations being with current tobacco smoking: yes, on most or all days (correlation=.114), difficulty not smoking for one day (correlation=.112), and smoking status: current (correlation=.108). 42.9% of sleep variables are associated with accelerated blood biochemistry aging, with the three largest associations being with sleeplessness/insomnia (correlation=.042), daytime dozing/sleeping (correlation=.035), and napping during the day (correlation=.034). 40.0% of sun exposure variables are associated with accelerated blood biochemistry aging, with the three largest associations being with time spent outdoors in winter (correlation=.069), time spent outdoors in summer (correlation=.059), and facial aging: older than you are (correlation=.034).

Conversely, the three environmental variable categories most associated with decelerated blood biochemistry aging are physical activity, smoking and sleep (Table S21). Specifically, 40.0% of physical activity variables are associated with decelerated blood biochemistry aging, with the three largest associations being with usual walking pace (correlation=.079), driving faster than motorway speed limit (correlation=.044), and duration walking for pleasure (correlation=.041). 29.2% of smoking variables are associated with decelerated blood biochemistry aging, with the three largest associations being with age started smoking (correlation=.115), time from waking to first cigarette (correlation=.113), and current tobacco smoking: no (correlation=.108). 28.6% of sleep variables are associated with decelerated blood biochemistry aging, with the two associations being with snoring (correlation=.034) and sleep duration (correlation=.022).

#### Socioeconomic variables associated with accelerated blood biochemistry aging

The three socioeconomic variable categories most associated with accelerated blood biochemistry aging are sociodemographics, education and employment (Table S23). Specifically, 57.1% of sociodemographic variables are associated with accelerated blood biochemistry aging, with the three largest associations being with private healthcare (correlation=.034), receiving attendance allowance (correlation=.022), and attendance/disability/mobility allowance: prefer not to answer (correlation=.013). 37.5% of education variables are associated with accelerated blood biochemistry aging, with the three largest associations being with qualifications: none of the qualifications listed (correlation=.066), qualifications: CSEs or equivalent (correlation=.027), and qualifications: prefer not to answer (correlation=.012). 18.2% of employment variables are associated with accelerated blood biochemistry aging, with the three largest associations being with having a job involving heavy manual or physical work (correlation=.042), mainly walking or standing (correlation=.032), and night shift work (correlation=.031).

Conversely, the three socioeconomic variable categories most associated with decelerated blood biochemistry aging are social support, education and household (Table S24). Specifically, 33.3% of social support variables are associated with decelerated blood biochemistry aging, with the three largest associations being with leisure/social activities: sports club or gym (correlation=.027), able to confide (correlation=.027), and frequency of friend/family visits (correlation=.018). 25.0% of education variables are associated with decelerated blood biochemistry aging, with the two associations being with having a college or university degree (correlation=.077) and having A/AS levels or equivalent (correlation=.048). 22.7% of household variables are associated with decelerated blood biochemistry aging, with the three largest associations being with income (correlation=.093), number of vehicles (correlation=.049) and number in household (correlation=.038).

### Correlation between blood cell, blood biochemistry and urine biochemistry aging

We estimated the phenotypic correlation between accelerated blood cell, blood biochemistry and urine biochemistry aging by computing the pairwise correlations between the three different dimensions of accelerated aging. Is a participant with accelerated blood biochemistry aging likely to also have accelerated blood cells aging? Accelerated blood cells and blood biochemistry aging are phenotypically .143+-.002 correlated, accelerated blood biochemistry and urine biochemistry are phenotypically .070+-.003 correlated, and blood cells and urine biochemistry are phenotypically .195+-.003 correlated.

Similarly, we estimated the genetic correlation between accelerated aging dimensions. Are the SNPs associated with accelerated aging in one dimension also associated with accelerated aging in another dimension? Accelerated blood cells and blood biochemistry aging are phenotypically .085+-.011 correlated, accelerated blood biochemistry and urine biochemistry are phenotypically −.080+-.043 correlated, and blood cells and urine biochemistry are phenotypically −.031+-.025 correlated.

Finally, we computed the correlations between the different accelerated aging dimensions in terms of their XWAS associations. For example, are the environmental variables associated with accelerated blood biochemistry aging also associated with accelerated blood cells aging? A summary of the all the correlations can be found in Table S25 and Figure S2. These results can be interactively explored at https://www.multidimensionality-of-aging.net/correlation_between_aging_dimensions/xwas_univariate.

### Predicting accelerated aging from biomarkers, clinical phenotypes, diseases, environmental variables and socioeconomic variables

We predicted accelerated aging in the three aging dimensions (blood biochemistry, blood cells, urine biochemistry) using variables from the different X-datasets categories (biomarkers, clinical phenotypes, diseases, environmental variables and socioeconomic variables). Specifically we built a model using the variables from each of their respective subcategories (e.g blood pressure biomarkers), and found that no dataset could explain more than 5% of the variance in accelerated aging.

## Discussion

### Blood biochemistry outperformed blood cells and urine biochemistry as an age predictor

Blood biochemistry biomarkers could explain approximately half the variance in chronological age (R^2^=48.6+-0.4%; RMSE=5.92+-0.02 years), in contrast to urine biochemistry and blood cells, which respectively explained 10.4+-0.3% and 12.5+-0.2% of the variance. The low performance of the urine biomarkers model can be explained by the limited number of biomarkers (four). It is likely that a model trained on a larger number of urine biomarkers would significantly outperform our current model in predicting age. In contrast, the low performance of blood cell biomarkers as age predictors is neither explained by the number of biomarkers (31), nor by the sample size (N=489,079). We conclude that, despite observable differences in normal blood count values throughout the different ages of life ^17^, blood count biomarkers can not, alone, predict age.

For the three aging dimensions, non-linear algorithms (GBM, neural network) outperformed the linear model (elastic net), suggesting that important information related to aging are encoded non-linearly and in complex interactions between the biomarkers.

### Comparison between our age predictors and the literature

We compare the performance of our models in predicting chronological age to models that were built on (1) laboratory biomarkers, which include all biomarkers listed under (2-4), (2) blood biochemistry biomarkers, (3) urine biochemistry biomarkers, and (4) blood count biomarkers.

#### Laboratory biomarkers

UKB’s laboratory biomarkers (blood count, blood biochemistry and urine biochemistry) have previously been analyzed to predict survival ^14,15^, but we are, to our knowledge, the first to leverage these to predict chronological age. We built separate models on blood biochemistry biomarkers, urine biochemistry biomarkers, and blood cells biomarkers (blood count), which complicates our task of comparing the performances of our models to the literature, as most publications did not make this distinction.

We briefly summarize the results of five studies which predicted chronological age using laboratory biomarkers (1-5). Three papers leveraged the NHANES ^18–25^ dataset to predict chronological age as a function of biochemical and blood count biomarkers ^6,7,26^. (1) Sagers et al. used a random forest to predict chronological age from 356 biomarkers collected from 67,563 participants aged 1-85 years. They obtained a R^2^ value of 92% and a MAE of 4.76 years ^6^. (2) Kendiukhov built a neural network on 926 predictors measured in 86,311 samples aged 10-85 years and predicted chronological age with a R^2^ of 78% and a MAE of 6.5 years ^26^. (3) Wood et al. used XGBoost ^27^ on 39 biomarkers collected on 46,739 participants aged 19-85 years. They predicted chronological age with a Pearson correlation of 0.75 for males and 0.77 in females ^7^, which is approximately equivalent to a R^2^ of 58%. (4) Putin et al. trained an ensemble of neural networks on 46 blood biomarkers from 62,419 blood biochemistry and cell count tests collected from patients aged 1-100 years and predicted chronological age with a R^2^ of 80% and a MAE of 6.07 years ^11^. (5) Wang et al. combined anthropometric measurements or vital signs with body fluid biomarkers. They leveraged 30 vital signs and 1,891 lab tests from 377,686 electronic medical records from patients of ages 18-85 to predict chronological age with a MAE of 7.58 years ^28^.

The diversity of the biomarkers covered by these models means that they should be compared to our models with caution; however, they still provide orders of magnitude for the prediction accuracy that can be reached when leveraging laboratory values to predict chronological age. Between the age predictors that we built, the model that is most similar to the aforementioned ones is our blood biochemistry-based model, which predicted chronological age with a R^2^ value of 48.6+-0.4% and a RMSE of 5.92+-0.02 years. We hypothesize that the underperformance of our model can be explained by (1) the narrower age range of the population in our study, (2) the greater the number of biomarkers included in the models found in the literature (n=46–1,921 compared to ours, 28) and (3) the types of biomarkers leveraged by the different publications.

#### Blood biochemistry

We predicted chronological age with a R^2^ value of 48.6±0.1% and a RMSE of 5.92±0.01 years using blood biochemistry biomarkers. Menni et al. predicted chronological age with a R^2^ of 59% using 22 blood metabolites collected on 6,055 participants aged 17-85 years ^12^. We hypothesize that the smaller R^2^ value of our blood biochemistry model (48.6±0.1%) can be explained by the larger age range of the cohort leveraged by Menni et al.

#### Urine biochemistry

We predicted chronological age with a R^2^ value of 10.4±0.1% and a RMSE of 7.72±0.01 using four urine biochemistry biomarkers. Hertel et al. used 59 urine nuclear magnetic resonance metabolomic biomarkers collected on 5,000 participants aged 20-79 years to predict chronological age with a R^2^ values of 56% for men and 61% for women (respective RMSEs: 11.2 and 10.37 years) ^9^. We hypothesize that our significantly smaller R^2^ value is driven by both the narrower age range covered by UKB and the smaller number of variables included in our model (four vs. 59). Our RMSE is smaller (7.72±0.01 years) than the RMSEs reported by Hertel et al., which could spuriously lead the reader to believe that our model performed better, but this is another consequence of UKB’s narrower age coverage.

#### Blood cells

We used blood count biomarkers to predict chronological age and obtained a R^2^ value of 12.5±0.01% and a RMSE of 7.59±0.01 years. To our knowledge, we are the first to predict chronological age from blood cell biomarkers. Alpert et al. built a predictor of survival based on the age trajectories of immune biomarkers (IMM-AGE) collected from 135 participants but did not directly predict chronological age ^29^. A scatter plot showing the correlation between IMM-AGE and chronological age in the Framingham Heart Study [FHS] dataset (age range 40-90 years) ^30^ can be found in Extended Data Fig.10b of their paper, but no correlation coefficient is reported. Both the correlation of our predictor and IMM-AGE with chronological age are modest (estimated R^2^=10±5% based on the figure). Because both models seem to be comparatively accurate despite IMM-AGE benefiting from the wider age range covered by the FHS dataset and considering that our model was specifically trained to predict chronological age, we hypothesize that our model slightly outperforms IMM-AGE as a chronological age predictor.

### Blood biochemistry, blood cells and urine biochemistry aging are three distinct phenotypes

Blood biochemistry, blood cells and urine biochemistry aging are three distinct phenotypes as shown by their weak phenotypic and genetic correlations. This low correlation can be explained in part by the low age prediction accuracy for the blood cell- and urine biochemistry-derived models. In contrast to these low correlations, we indeed observed that accelerated blood biochemistry aging is associated with blood cells and urine biochemistry biomarkers. More generally, in terms of nongenetic correlations (similarity in terms of associations with biomarkers, phenotypes, diseases, environmental and socioeconomic variables), we found that blood biochemistry and blood cells accelerated aging are correlated (e.g they are influenced similarly by environmental factors), whereas urine biochemistry accelerated aging stands apart and is negatively correlated with the two other accelerated aging phenotypes.

### Blood and urine features driving age prediction

The biomarkers selected as most important by the different models undergo important changes with age, as observed by their pairwise correlation with the target variable, and as reported in the literature.

#### Blood biochemistry

The biomarkers prioritized by the models are involved in sexual, metabolic, kidney and liver functions. Sex hormone binding globulin and testosterone levels decrease with age in men ^31–33^. Glycated hemoglobin, apolipoprotein B, LDL and IGF-1 are involved in metabolism. Specifically, glycated hemoglobin is a marker of glycemic control and diabetes that increases with age ^34^, and apolipoprotein B is a protein involved in the transport of cholesterol and a biomarker for atherosclerosis whose levels increase with age ^35^. Insulin-like growth factor 1 is an anabolic hormone involved in growth whose serum levels decrease with age ^36^, and LDL levels decrease with age ^37^. Cystatin C, serum creatinine and serum urea are three kidney function biomarkers whose levels increase with age ^38–40^. Alanine aminotransferase is a marker of liver injury that decreases with age ^41^.

#### Urine biochemistry

Urine creatinine is a biomarker of kidney function which decreases with age as creatinine clearance declines ^42^. Similarly, sodium in urine decreases with age as the kidney becomes less able to remove excess sodium ^43^. Elevated microalbumin levels (microalbuminuria) and an elevated ratio between microalbumin and creatinine levels are a sign of poor renal function and of vascular damage ^44^.

#### Blood cells

According to the literature red blood cell distribution width ^45^ and mean corpuscular volume ^46^ increase with age, whereas red blood cell count ^47^, platelet crit ^48^, lymphocyte percentage ^49^ and hemoglobin levels ^50^ decrease with age. Mean sphered volume, mean reticulocyte volume and neutrophil count are positively correlated with age in UKB’s cohort (respective correlations: .107, .099 and .022). These observations explain the importance of the features selected by the models.

### Accelerated aging is partially heritable

We found that accelerated aging was 26.2-10.5% heritable, depending on the aging dimension. The SNPs highlighted by the GWAS reflect, as expected, the feature importances of the models. For example, the GWAS we performed on accelerated blood biochemistry aging identified the genes encoding *SHBG* and cystatin C as significantly associated with these aging phenotypes. This finding is intuitive considering that these two biomarkers were respectively ranked as the first and the sixth most important predictors by the model we trained to predict chronological age as a function of blood biochemistry biomarkers. *SHBG* and cystatin C levels increase with age (respective correlations of .06±.01 and .30±.02) so accelerated biochemistry agers are likely to have higher *SHBG* and cystatin C levels than an average person of the same chronological age. Genes for which some alleles are associated with a higher/lower serum SHBG and cystatin C levels are therefore likely to be associated with accelerated/decelerated biochemistry aging by the GWAS. Similarly, the model ranked glycosylated hemoglobin [HbA1c] as the third most important predictor and glucose as the 19th most important predictors for chronological, which possibly explains why the GWAS identified *MPDU1*, *HK1* and *G6PC2* as significantly associated with accelerated aging, since mutations in *MPDU1* lead to glycosylation disorders ^16^ and both *HK1* and *G6PC2* are enzymes involved in the glucose pathway ^16^. The GWAS also highlighted UDP glucuronosyltransferase genes, which are linked to liver function and more specifically to drug metabolism ^16^. Deficiency in UDP glucuronosyltransferase can lead to increased drug hepatotoxicity, which increases alanine aminotransferase [ALT] levels ^51^. ALT is a biomarker for liver damage and was ranked as the tenth most important predictor for chronological age by our model. Finally, the GWAS highlighted *CETP* (involved in cholesterol metabolism) and *ABCB11* (involved in statins metabolism, which regulate cholesterol level), which can be explained by the blood biochemistry model, which included cholesterol, LDL, HDL and triglycerides as predictors (respectively ranked as 18th, seventh, 21th and 17th most important predictors).

Similarly, for blood cell aging, the four most important age predictors are red blood cell distribution width, red blood cell count, mean corpuscular volume and mean sphered cell volume. The GWAS highlighted several genes related to red blood cells such as *HBS1L* (involved in erythrocyte volume, hemoglobin content, erythrocyte count, platelet count and monocyte count, and linked to sickle cell disease and beta-thalassemia), *SPTA1* (a component of red blood cell plasma membrane and linked to erythrocyte diseases such as elliptocytosis 2, pyropoikilocytosis, and type 3 spherocytosis), and *ANK1* (linked to spherocytosis). The 5th most important predictor for age is platelet crit, and the GWAS highlighted *TUBB1* (expressed in thrombocytes and megakaryocytes and linked to macrothrombocytopenia) and *WDR66* (involved in mean platelet volume).

We emphasize, however, that the interpretation of these loci are different than the associations uncovered in the GWASs of the individual subdimension traits themselves (e.g., *SHBG* and creatinine). Specifically, SNPs and loci uncovered in our study may hypothetically exert their effect, if causal, along the axis of biological age in a pleiotropic manner.

### Accelerated blood biochemistry aging is linked to aging in other organ systems

Accelerated blood biochemistry aging is associated with cardiovascular biomarkers (e.g pulse rate, arterial stiffness, blood pressure, waist circumference, GMI, hip circumference), as well as with lung function, ocular health (eyesight, intraocular pressure), hearing acuity, musculoskeletal health (hand grip strength, bone density), brain anatomical features and cognitive function biomarkers. Similarly it is associated with clinical phenotypes in diverse organ systems, such as chest pain, breathing issues, claudication, joint pain, mental health, hearing, eyesight and oral health. Interestingly, blood aging is correlated with facial aging. Finally, it is associated with diverse diseases, such as cardiovascular disease, gastrointestinal disorders, poor general health, respiratory diseases, kidney disease, metabolic disorders, neuropathies, mental diseases, skin diseases, eye diseases, fractures, cancer, infections, and others. These findings suggest that blood aging is linked to aging in other organ systems, an hypothesis which we further investigate in a separate paper ^52^.

### Environmental and social and economic factors associated with accelerated blood biochemistry aging

In terms of environmental variables, accelerated blood biochemistry aging is associated with smoking (including maternal smoking around birth), poor sleep and sedentarity. Decelerated blood biochemistry aging, in contrast, is associated with physical activity. These findings are aligned with the literature ^53–55^. Some diet factors are associated with accelerated aging (e.g white bread, high sugar cereals), while others are associated with decelerated aging (whole grain bread, oily fish). Alcohol intake showed mixed associations, with alcohol intake frequency being associated with accelerated aging, whereas usually taking alcohol with meals and red wine intake are associated with decelerated aging, reflecting a complex literature ^56^.

In terms of socioeconomic status, being wealthy, education and emotionally supported (e.g able to confide, frequency of friend/family visits) were associated with decelerated blood biochemistry aging. Social support is associated with better health ^57^, and education might improve aging via health literacy ^58^. The correlation between wealth and life expectancy is also strong, with the richest 1% US males and females respectively living 14.6±0.2 and 10.1±0.2 years longer than their poorest 1% counterparts ^59^.

### Limitations

We identify three key limitations to our work. First, the age range covered by the UKB is narrow (37-82 years). Our models therefore do not inform about the aging process during childhood and young adulthood and centenarian years, and would likely poorly generalize on these populations. Second, the poor predictive performance of blood count and urine biochemistry biomarkers as chronological age predictors, in part due to the limited number of urine biomarkers, limits the power of the subsequent steps of our analysis (e.g GWAS, XWAS, correlation between aging dimensions). Third, UKB is an observational cohort and a largely cross-sectional dataset. As a consequence, the correlations we report between accelerated aging and other factors (biomarkers, phenotypes, diseases, environmental and socioeconomic variables) do not allow us to infer causation.

## Methods

### Data and materials availability

We used the UK Biobank (project ID: 52887). The code can be found at https://github.com/Deep-Learning-and-Aging. The results can be interactively and extensively explored at https://www.multidimensionality-of-aging.net/. We will make the biological age phenotypes available through UK Biobank upon publication. The GWAS results can be found at https://www.dropbox.com/s/59e9ojl3wu8qie9/Multidimensionality_of_aging-GWAS_results.zip?dl=0.

### Software

Our code can be found at https://github.com/Deep-Learning-and-Aging. For the genetics analysis, we used the BOLT-LMM ^60,61^ and BOLT-REML ^62^ softwares. We coded the parallel submission of the jobs in Bash ^63^.

### Cohort Dataset: Participants of the UK Biobank

We leveraged the UK Biobank^13^ cohort (project ID: 52887). The UKB cohort consists of data originating from a large biobank collected from 502,211 de-identified participants in the United Kingdom that were aged between 37 years and 74 years at enrollment (starting in 2006). The Harvard internal review board (IRB) deemed the research as non-human subjects research (IRB: IRB16-2145).

### Data types and Preprocessing

The data preprocessing step is different for demographic variables and biomarkers. For demographics variables, first, we removed out the UKB samples for which age or sex was missing. For sex, we used the genetic sex when available, and the self-reported sex when genetic sex was not available. We computed age as the difference between the date when the participant attended the assessment center and the year and month of birth of the participant to estimate the participant’s age with greater precision. We one-hot encoded ethnicity. For the biomarkers, we did not perform any preprocessing, aside from the normalization that is described under cross-validation further below. The complete list of biomarkers can be found in Table S7 under “BloodBiochemistry”, “BloodCells” and “UrineBiochemistry”.

### Machine learning algorithms

We used three different algorithms to predict age. Elastic Nets [EN] (a regularized linear regression that represents a compromise between ridge regularization and LASSO regularization), Gradient Boosted Machines [GBM] (LightGBM implementation ^64^), and Neural Networks [NN]. The choice of these three algorithms represents a compromise between interpretability and performance. Linear regressions and their regularized forms (LASSO ^65^, ridge ^66^, elastic net ^67^) are highly interpretable using the regression coefficients but are poorly suited to leverage non-linear relationships or interactions between the features and therefore tend to underperform compared to the other algorithms. In contrast, neural networks ^68,69^ are complex models, which are designed to capture non-linear relationships and interactions between the variables. However, tools to interpret them are limited ^70^ so they are closer to a “black box”. Tree-based methods such as random forests ^71^, gradient boosted machines ^72^ or XGBoost ^27^ represent a compromise between linear regressions and neural networks in terms of interpretability. They tend to perform similarly to neural networks when limited data is available, and the feature importances can still be used to identify which predictors played an important role in generating the predictions. However, unlike linear regression, feature importances are always non-negative values, so one cannot interpret whether a predictor is associated with older or younger age. We also performed preliminary analyses with other tree-based algorithms, such as random forests ^71^, vanilla gradient boosted machines ^72^ and XGBoost ^27^. We found that they performed similarly to LightGBM, so we only used this last algorithm as a representative for tree-based algorithms in our final calculations.

### Training, tuning and predictions

We split the entire dataset into ten data folds and performed a nested-cross validation. We describe the splitting of the data into different folds and the tuning procedures in greater detail in the Supplementary.

### Interpretability of the machine learning predictions

To interpret the models, we used the regression coefficients for the elastic nets, the feature importances for the GBMs, and a permutation test for the fully connected neural networks (Supplementary Methods).

### Ensembling to improve prediction and define aging dimensions

For each dataset, we ensembled the three models obtained from the elastic net, the GBM and the neural network. We built each ensemble model separately on each of the ten data folds. For example, to build the ensemble model on the testing predictions of the data fold #1, we trained and tuned an elastic net on the validation predictions from the data fold #0 using a 10-folds inner cross-validation, as the validation predictions on fold #0 and the testing predictions on fold #1 are generated by the same model. We used the same hyperparameters space and Bayesian hyperparameters optimization method as we did for the inner cross-validation we performed during the tuning of the non-ensemble models.

To summarize, the testing ensemble predictions are computed by concatenating the testing predictions generated by ten different elastic nets, each of which was trained and tuned using a 10-folds inner cross-validation on one validation data fold (10% of the full dataset) and tested on one testing fold. This is different from the inner-cross validation performed when training the non-ensemble models, which was performed on the “training+validation” data folds, so on 9 data folds (90% of the dataset).

### Evaluating the performance of models

We evaluated the performance of the models using two different metrics: R-Squared [R^2^] and root mean squared error [RMSE]. We computed a confidence interval on the performance metrics in two different ways. First, we computed the standard deviation between the different data folds. The test predictions on each of the ten data folds are generated by ten different models, so this measure of standard deviation captures both model variability and the variability in prediction accuracy between samples. Second, we computed the standard deviation by bootstrapping the computation of the performance metrics 1,000 times. This second measure of variation does not capture model variability but evaluates the variance in the prediction accuracy between samples.

### Biological age definition

We defined the biological age of participants as the prediction generated by the model corresponding to aging dimension, after correcting for the bias in the residuals. We indeed observed a bias in the residuals. For each model, participants on the older end of the chronological age distribution tend to be predicted younger than they are. Symmetrically, participants on the younger end of the chronological age distribution tend to be predicted older than they are. This bias does not seem to be biologically driven. Rather it seems to be statistically driven, as the same 60-year-old individual will tend to be predicted younger in a cohort with an age range of 60-80 years, and to be predicted older in a cohort with an age range of 60-80. We ran a linear regression on the residuals as a function of age for each model and used it to correct each prediction for this statistical bias.

After defining biological age as the corrected prediction, we defined accelerated aging as the corrected residuals. For example, a 60-year-old whose blood biochemistry biomarkers predicted an age of 70 years old after correction for the bias in the residuals is estimated to have a blood biochemistry age of 70 years, and an accelerated blood biochemistry aging of ten years.

It is important to understand that this step of correction of the predictions and the residuals takes place after the evaluation of the performance of the models but precedes the analysis of the biological ages properties.

### Genome-wide association study of accelerated aging

The UKB contains genome-wide genetic data for 488,251 of the 502,492 participants^73^ under the hg19/GRCh37 build. We used the average accelerated aging value over the different samples collected over time (see Supplementary - Generating average predictions for each participant). Next, we performed genome wide association studies [GWASs] to identify single-nucleotide polymorphisms [SNPs] associated with accelerated aging in each aging dimension using BOLT-LMM ^60,61^ and estimated the SNP-based heritability for each of our biological age phenotypes, and we computed the genetic pairwise correlations between dimensions using BOLT-REML ^62^. We used the v3 imputed genetic data to increase the power of the GWAS, and we corrected all of them for the following covariates: age, sex, ethnicity, the assessment center that the participant attended when their DNA was collected, and the 20 genetic principal components precomputed by the UKB. We used the linkage disequilibrium [LD] scores from the 1,000 Human Genomes Project ^74^. To avoid population stratification, we performed our GWAS on individuals with White ethnicity.

#### Identification of SNPs associated with accelerated aging

We identified the SNPs associated with accelerated aging dimensions using the BOLT-LMM ^60,61^ software (p-value of 5e-8). The sample size for the genotyping of the X chromosome is one thousand samples smaller than for the autosomal chromosomes. We therefore performed two GWASs for each aging dimension. (1) excluding the X chromosome, to leverage the full autosomal sample size when identifying the SNPs on the autosome. (2) including the X chromosome, to identify the SNPs on this sex chromosome. We then concatenated the results from the two GWASs to cover the entire genome, at the exception of the Y chromosome. We plotted the results using a Manhattan plot and a volcano plot. We used the bioinfokit ^75^ python package to generate the Manhattan plots. We generated quantile-quantile plots [Q-Q plots] to estimate the p-value inflation as well.

#### Heritability and genetic correlation

We estimated the heritability of the accelerated aging dimensions using the BOLT-REML ^62^ software. We included the X chromosome in the analysis and corrected for the same covariates as we did for the GWAS. Using the same software and parameters, we computed the genetic correlations between accelerated aging in the different aging dimensions. We annotated the significant SNPs with their matching genes using the following four steps pipeline. (1) We annotated the SNPs based on the rs number using SNPnexus ^76–80^. When the SNP was between two genes, we annotated it with the nearest gene. (2) We used SNPnexus to annotate the SNPs that did not match during the first step, this time using their genomic coordinates. After these two first steps, 30 out of the 9,697 significant SNPs did not find a match. (3) We annotated these SNPs using LocusZoom ^81^. Unlike SNPnexus, LocusZoom does not provide the gene types, so we filled this information with GeneCards ^16^. After this third step, four genes were not matched. (4) We used RCSB Protein Data Bank ^82^ to annotate three of the four missing genes.

### Non-genetic correlates of accelerated aging

We identified non-genetically measured (i.e factors not measured on a GWAS array) correlates of each aging dimension, which we classified in six categories: biomarkers, clinical phenotypes, diseases, family history, environmental, and socioeconomic variables. We refer to the union of these association analyses as an X-Wide Association Study [XWAS]. (1) We define as biomarkers the scalar variables measured on the participant, which we initially leveraged to predict age (e.g. blood pressure, Table S7). (2) We define clinical phenotypes as other biological factors not directly measured on the participant, but instead collected by the questionnaire, and which we did not use to predict chronological age. For example, one of the clinical phenotypes categories is eyesight, which contains variables such as “wears glasses or contact lenses”, which is different from the direct refractive error measurements performed on the participants, which are considered “biomarkers” (Table S10). (3) Diseases include the different medical diagnoses categories listed by UKB (Table S13). (4) Family history variables include illnesses of family members (Table S16). (5) Environmental variables include alcohol, diet, electronic devices, medication, sun exposure, early life factors, medication, sun exposure, sleep, smoking, and physical activity variables collected from the questionnaire (Table S19). (6) Socioeconomic variables include education, employment, household, social support and other sociodemographics (Table S22). We provide information about the preprocessing of the XWAS in the Supplementary Methods.

## Data Availability

https://www.multidimensionality-of-aging.net/

https://www.dropbox.com/s/59e9ojl3wu8qie9/Multidimensionality_of_aging-GWAS_results.zip?dl=0

## Author Contributions

**Alan Le Goallec:** (1) Designed the project. (2) Supervised the project. (3) Ensembled the models, evaluated their performance, computed biological ages and estimated the correlation structure between the aging dimensions. (4) Performed the genome wide association studies. (5) Designed the website. (6) Wrote the manuscript.

**Samuel Diai:** (1) Predicted chronological age from the scalar features. (2) Coded the algorithm to obtain balanced data folds across the different datasets. (3) Wrote the python class to build an ensemble model using a cross-validated elastic net. (4) Performed the X-wide association study. (5) Implemented a first version of the website https://www.multidimensionality-of-aging.net/.

**Théo Vincent:** (1) Website data engineer. (2) Implemented a second version of the website https://www.multidimensionality-of-aging.net/.

**Chirag J. Patel:** (1) Supervised the project. (2) Edited the manuscript. (3) Provided funding.

## Acknowledgments

We would like to thank Raffaele Potami from Harvard Medical School research computing group for helping us utilize O2’s computing resources. We thank HMS RC for computing support. We also want to acknowledge UK Biobank for providing us with access to the data they collected. The UK Biobank project number is 52887.

## Funding

NIEHS R00 ES023504

NIEHS R21 ES25052.

NIAID R01 AI127250

NSF 163870

MassCATS, Massachusetts Life Science Center

Sanofi

The funders had no role in the study design or drafting of the manuscript(s).

## References

1. Fuster, V. Changing Demographics: A New Approach to Global Health Care Due to the Aging Population. J. Am. Coll. Cardiol. 69, 3002–3005 (2017).

2. Kennedy, B. K. et al. Geroscience: linking aging to chronic disease. Cell 159, 709–713 (2014).

3. Johnson, N. B., Hayes, L. D., Brown, K., Hoo, E. C. & Ethier, K. A. CDC National Health Report: leading causes of morbidity and mortality and associated behavioral risk and protective factors—United States, 2005--2013. (2014).

4. Jylhävä, J., Pedersen, N. L. & Hägg, S. Biological Age Predictors. EBioMedicine 21, 29–36 (2017).

5. Zhavoronkov, A., Li, R., Ma, C. & Mamoshina, P. Deep biomarkers of aging and longevity: from research to applications. Aging 11, (2019).

6. Sagers, L., Melas-Kyriazi, L., Patel, C. J. & Manrai, A. K. Prediction of chronological and biological age from laboratory data. Aging 12, 7626–7638 (2020).

7. Wood, T., Kelly, C., Roberts, M. & Walsh, B. An interpretable machine learning model of biological age. F1000Res. 8, (2019).

8. Mamoshina, P. et al. Population Specific Biomarkers of Human Aging: A Big Data Study Using South Korean, Canadian, and Eastern European Patient Populations. J. Gerontol. A Biol. Sci. Med. Sci. 73, 1482–1490 (2018).

9. Hertel, J. et al. Measuring Biological Age via Metabonomics: The Metabolic Age Score. J. Proteome Res. 15, 400–410 (2016).

10. van den Akker, E. B. et al. Metabolic Age Based on the BBMRI-NL 1 H-NMR Metabolomics Repository as Biomarker of Age-related Disease. Circulation: Genomic and Precision Medicine (2020) doi:10.1161/circgen.119.002610.

11. Putin, E. et al. Deep biomarkers of human aging: Application of deep neural networks to biomarker development. Aging 8, 1021–1033 (2016).

12. Menni, C. et al. Metabolomic markers reveal novel pathways of ageing and early development in human populations. Int. J. Epidemiol. 42, 1111–1119 (2013).

13. Sudlow, C. et al. UK biobank: an open access resource for identifying the causes of a wide range of complex diseases of middle and old age. PLoS Med. 12, e1001779 (2015).

14. Chan, M. S. et al. Biological age in UK Biobank: biomarker composition and prediction of mortality, coronary heart disease and hospital admissions. doi:10.1101/2019.12.12.19014720.

15. Ganna, A. & Ingelsson, E. 5 year mortality predictors in 498,103 UK Biobank participants: a prospective population-based study. Lancet 386, 533–540 (2015).

16. Stelzer, G. et al. The GeneCards Suite: From Gene Data Mining to Disease Genome Sequence Analyses. Curr. Protoc. Bioinformatics 54, 1.30.1–1.30.33 (2016).

17. Zierk, J. et al. Blood counts in adult and elderly individuals: defining the norms over eight decades of life. Br. J. Haematol. 189, 777–789 (2020).

18. Statistics, U. S. D. of H. A. H. S. C. F. D. C. A. P. N. C. F. H. & United States Department of Health and Human Services. Centers for Disease Control and Prevention. National Center for Health Statistics. National Health and Nutrition Examination Survey (NHANES), 1999-2000. ICPSR Data Holdings (2009) doi:10.3886/icpsr25501.v3.

19. Statistics, U. S. D. of H. A. H. S. C. F. D. C. A. P. N. C. F. H. & United States Department of Health and Human Services. Centers for Disease Control and Prevention. National Center for Health Statistics. National Health and Nutrition Examination Survey (NHANES), 2001-2002. ICPSR Data Holdings (2010) doi:10.3886/icpsr25502.v4.

20. Statistics, U. S. D. of H. A. H. S. C. F. D. C. A. P. N. C. F. H. & United States Department of Health and Human Services. Centers for Disease Control and Prevention. National Center for Health Statistics. National Health and Nutrition Examination Survey (NHANES), 2003-2004. ICPSR Data Holdings (2010) doi:10.3886/icpsr25503.v5.

21. Statistics, U. S. D. of H. A. H. S. C. F. D. C. A. P. N. C. F. H. & United States Department of Health and Human Services. Centers for Disease Control and Prevention. National Center for Health Statistics. National Health and Nutrition Examination Survey (NHANES), 2005-2006. ICPSR Data Holdings (2010) doi:10.3886/icpsr25504.v3.

22. Statistics, U. S. D. of H. A. H. S. C. F. D. C. A. P. N. C. F. H. & United States Department of Health and Human Services. Centers for Disease Control and Prevention. National Center for Health Statistics. National Health and Nutrition Examination Survey (NHANES), 2007-2008. ICPSR Data Holdings (2010) doi:10.3886/icpsr25505.v2.

23. for Disease Control, C., Prevention & Others. National Health and Nutrition Examination Survey. 2010. (2010).

24. Johnson, C. L., Dohrmann, S. M., Burt, V. L. & Mohadjer, L. K. National health and nutrition examination survey: sample design, 2011-2014. Vital Health Stat. 21–33 (2014).

25. for Disease Control, C. & Prevention. National health and nutrition examination survey. 2020.

26. Kendiukhov, I. AI-based investigation of molecular biomarkers of longevity. Biogerontology (2020) doi:10.1007/s10522-020-09890-y.

27. Chen, T. & Guestrin, C. XGBoost: A Scalable Tree Boosting System. in Proceedings of the 22nd ACM SIGKDD International Conference on Knowledge Discovery and Data Mining 785–794 (Association for Computing Machinery, 2016).

28. Wang, Z. et al. Predicting age by mining electronic medical records with deep learning characterizes differences between chronological and physiological age. J. Biomed. Inform. 76, 59–68 (2017).

29. Alpert, A. et al. A clinically meaningful metric of immune age derived from high-dimensional longitudinal monitoring. Nat. Med. 25, 487–495 (2019).

30. D’Agostino, R. B. et al. General cardiovascular risk profile for use in primary care. Circulation 117, 743–753 (2008).

31. Feldman, H. A. et al. Age trends in the level of serum testosterone and other hormones in middle-aged men: longitudinal results from the Massachusetts male aging study. J. Clin. Endocrinol. Metab. 87, 589–598 (2002).

32. Sowers, M. F., Beebe, J. L., McConnell, D., Randolph, J. & Jannausch, M. Testosterone Concentrations in Women Aged 25-50 Years: Associations with Lifestyle, Body Composition, and Ovarian Status. Am. J. Epidemiol. 153, 256–264 (2001).

33. Maggio, M. et al. Sex hormone binding globulin levels across the adult lifespan in women — The role of body mass index and fasting insulin. Journal of Endocrinological Investigation vol. 31 597–601 (2008).

34. Suvarna H.I, S., Moodithaya, S. & Sharma, R. Metabolic and Cardiovascular Ageing Indices in Relation to Glycated Haemoglobin in Healthy and Diabetic Subjects. Curr. Aging Sci. 10, 201–210 (2017).

35. Schaefer, E. J. et al. Effects of age, gender, and menopausal status on plasma low density lipoprotein cholesterol and apolipoprotein B levels in the Framingham Offspring Study. J. Lipid Res. 35, 779–792 (1994).

36. Zhu, H. et al. Reference ranges for serum insulin-like growth factor I (IGF-I) in healthy Chinese adults. PLoS One 12, e0185561 (2017).

37. Ferrara, A., Barrett-Connor, E. & Shan, J. Total, LDL, and HDL cholesterol decrease with age in older men and women. The Rancho Bernardo Study 1984-1994. Circulation 96, 37–43 (1997).

38. Odden, M. C. et al. Age and cystatin C in healthy adults: a collaborative study. Nephrol. Dial. Transplant 25, 463–469 (2010).

39. Tiao, J. Y.-H., Semmens, J. B., Masarei, J. R. L. & Lawrence-Brown, M. M. D. The effect of age on serum creatinine levels in an aging population: relevance to vascular surgery. Cardiovasc. Surg. 10, 445–451 (2002).

40. Musch, W., Verfaillie, L. & Decaux, G. Age-Related Increase in Plasma Urea Level and Decrease in Fractional Urea Excretion: Clinical Application in the Syndrome of Inappropriate Secretion of Antidiuretic Hormone. Clinical Journal of the American Society of Nephrology vol. 1 909–914 (2006).

41. Dong, M. H., Bettencourt, R., Barrett-Connor, E. & Loomba, R. Alanine aminotransferase decreases with age: the Rancho Bernardo Study. PLoS One 5, e14254 (2010).

42. Rowe, J. W., Andres, R., Tobin, J. D., Norris, A. H. & Shock, N. W. The effect of age on creatinine clearance in men: a cross-sectional and longitudinal study. J. Gerontol. 31, 155–163 (1976).

43. Schmidt, R. J., Beierwaltes, W. H. & Baylis, C. Effects of aging and alterations in dietary sodium intake on total nitric oxide production. Am. J. Kidney Dis. 37, 900–908 (2001).

44. Chowta, N. K., Pant, P. & Chowta, M. N. Microalbuminuria in diabetes mellitus: Association with age, sex, weight, and creatinine clearance. Indian J. Nephrol. 19, 53–56 (2009).

45. Hoffmann, J. J. M. L., Nabbe, K. C. A. M. & van den Broek, N. M. A. Effect of age and gender on reference intervals of red blood cell distribution width (RDW) and mean red cell volume (MCV). Clin. Chem. Lab. Med. 53, 2015–2019 (2015).

46. Gamaldo, A. A., Ferrucci, L., Rifkind, J. M. & Zonderman, A. B. Age-related changes in mean corpuscular volume in adult whites and African Americans. J. Am. Geriatr. Soc. 59, 1763 (2011).

47. Kubota K. et al. Changes in the blood cell counts with aging. Nihon Ronen Igakkai Zasshi 28, 509–514 (1991).

48. Santimone, I. et al. White blood cell count, sex and age are major determinants of heterogeneity of platelet indices in an adult general population: results from the MOLI-SANI project. Haematologica 96, 1180–1188 (2011).

49. Shahabuddin, S., Al-Ayed, I., Gad El-Rab, M. O. & Qureshi, M. I. Age-Related Changes in Blood Lymphocyte Subsets of Saudi Arabian Healthy Children. Clinical Diagnostic Laboratory Immunology vol. 7 990–990 (2000).

50. Hawkins, W. W., Speck, E. & Leonard, V. G. Variation of the hemoglobin level with age and sex. Blood 9, 999–1007 (1954).

51. De Morais, S. M. & Wells, P. G. Deficiency in bilirubin UDP-glucuronyl transferase as a genetic determinant of acetaminophen toxicity. J. Pharmacol. Exp. Ther. 247, 323–331 (1988).

52. Le Goallec, A. et al. Analyzing the multidimensionality of biological aging with the tools of deep learning across diverse image-based and physiological indicators yields robust age predictors. medRxiv (2021).

53. Warburton, D. E. R., Nicol, C. W. & Bredin, S. S. D. Health benefits of physical activity: the evidence. CMAJ 174, 801–809 (2006).

54. Hale, L., Troxel, W. & Buysse, D. J. Sleep Health: An Opportunity for Public Health to Address Health Equity. Annu. Rev. Public Health 41, 81–99 (2020).

55. Jha, P. The hazards of smoking and the benefits of cessation: a critical summation of the epidemiological evidence in high-income countries. Elife 9, (2020).

56. Burton, R. & Sheron, N. No level of alcohol consumption improves health. The Lancet vol. 392 987–988 (2018).

57. Reblin, M. & Uchino, B. N. Social and emotional support and its implication for health. Curr. Opin. Psychiatry 21, 201–205 (2008).

58. Liu, C. et al. What is the meaning of health literacy? A systematic review and qualitative synthesis. Family medicine and community health 8, (2020).

59. Chetty, R. et al. The Association Between Income and Life Expectancy in the United States, 2001-2014. JAMA 315, 1750–1766 (2016).

60. Loh, P.-R. et al. Efficient Bayesian mixed-model analysis increases association power in large cohorts. Nat. Genet. 47, 284–290 (2015).

61. Loh, P.-R., Kichaev, G., Gazal, S., Schoech, A. P. & Price, A. L. Mixed-model association for biobank-scale datasets. Nat. Genet. 50, 906–908 (2018).

62. Loh, P.-R. et al. Contrasting genetic architectures of schizophrenia and other complex diseases using fast variance-components analysis. Nature Genetics vol. 47 1385–1392 (2015).

63. Gnu, P. Free Software Foundation. Bash (3. 2. 48)[Unix shell program] (2007).

64. Ke, G. et al. LightGBM: A Highly Efficient Gradient Boosting Decision Tree. in Advances in Neural Information Processing Systems 30 (eds. Guyon, I. et al.) 3146–3154 (Curran Associates, Inc., 2017).

65. Tibshirani, R. Regression shrinkage and selection via the lasso. J. R. Stat. Soc. Series B Stat. Methodol. 58, 267–288 (1996).

66. Hoerl, A. E. & Kennard, R. W. Ridge Regression: Biased Estimation for Nonorthogonal Problems. null 12, 55–67 (1970).

67. Zou, H. & Hastie, T. Regularization and variable selection via the elastic net. J. R. Stat. Soc. Series B Stat. Methodol. 67, 301–320 (2005).

68. Rosenblatt, F. The Perceptron: A Theory of Statistical Separability in Cognitive Systems (Project Para). (Cornell Aeronautical Laboratory, 1958).

69. Popescu, M.-C., Balas, V. E., Perescu-Popescu, L. & Mastorakis, N. Multilayer perceptron and neural networks. WSEAS Trans. Circuits and Syst. 8, (2009).

70. Ribeiro, M. T., Singh, S. & Guestrin, C. ‘Why should I trust you?’ Explaining the predictions of any classifier. in Proceedings of the 22nd ACM SIGKDD international conference on knowledge discovery and data mining 1135–1144 (2016).

71. Breiman, L. Random Forests. Mach. Learn. 45, 5–32 (2001).

72. Friedman, J. H. Greedy Function Approximation: A Gradient Boosting Machine. Ann. Stat. 29, 1189–1232 (2001).

73. Bycroft, C. et al. Genome-wide genetic data on\ 500,000 UK Biobank participants. BioRxiv 166298 (2017).

74. Consortium, T. 1000 G. P. & The 1000 Genomes Project Consortium. A global reference for human genetic variation. Nature vol. 526 68–74 (2015).

75. Bedre, R. reneshbedre/bioinfokit: Bioinformatics data analysis and visualization toolkit. (2020). doi:10.5281/zenodo.3965241.

76. Oscanoa, J. et al. SNPnexus: a web server for functional annotation of human genome sequence variation (2020 update). Nucleic Acids Res. 48, W185–W192 (2020).

77. Dayem Ullah, A. Z., et al. SNPnexus: assessing the functional relevance of genetic variation to facilitate the promise of precision medicine. Nucleic Acids Res. 46, W109–W113 (2018).

78. Ullah, A. Z. D., Dayem Ullah, A. Z., Lemoine, N. R. & Chelala, C. A practical guide for the functional annotation of genetic variations using SNPnexus. Briefings in Bioinformatics vol. 14 437–447 (2013).

79. Dayem Ullah, A. Z., Lemoine, N. R. & Chelala, C. SNPnexus: a web server for functional annotation of novel and publicly known genetic variants (2012 update). Nucleic Acids Res. 40, W65–70 (2012).

80. Chelala, C., Khan, A. & Lemoine, N. R. SNPnexus: a web database for functional annotation of newly discovered and public domain single nucleotide polymorphisms. Bioinformatics 25, 655–661 (2009).

81. Pruim, R. J. et al. LocusZoom: regional visualization of genome-wide association scan results. Bioinformatics 26, 2336–2337 (2010).

82. Berman, H. M. et al. The protein data bank. Nucleic Acids Res. 28, 235–242 (2000).

83. Van Rossum, G. & Drake, F. L. The Python Language Reference Manual. (Network Theory Limited, 2011).

84. Oliphant, T. E. A guide to NumPy. vol. 1 (Trelgol Publishing USA, 2006).

85. Walt, S. van der, van der Walt, S., Chris Colbert, S. & Varoquaux, G. The NumPy Array: A Structure for Efficient Numerical Computation. Computing in Science & Engineering vol. 13 22–30 (2011).

86. McKinney, W. & Others. Data structures for statistical computing in python. in Proceedings of the 9th Python in Science Conference vol. 445 51–56 (Austin, TX, 2010).

87. Hunter, J. D. Matplotlib: A 2D Graphics Environment. Comput. Sci. Eng. 9, 90–95 (2007).

88. Inc, P. T. Collaborative data science. Montreal: Plotly Technologies Inc Montral (2015).

89. Clark, A. Pillow Python Imaging Library. Pillow—Pillow (PIL Fork) 5. 4. 1 documentation (2018).

90. Virtanen, P. et al. SciPy 1.0: fundamental algorithms for scientific computing in Python. Nature Methods vol. 17 261–272 (2020).

91. Oliphant, T. E. Python for Scientific Computing. Computing in Science Engineering 9, 10–20 (2007).

92. Millman, K. J., Jarrod Millman, K. & Aivazis, M. Python for Scientists and Engineers. Computing in Science & Engineering vol. 13 9–12 (2011).

93. Pedregosa, F. et al. Scikit-learn: Machine learning in Python. the Journal of machine Learning research 12, 2825–2830 (2011).

94. Bergstra, J., Yamins, D. & Cox, D. D. Hyperopt: A python library for optimizing the hyperparameters of machine learning algorithms. in Proceedings of the 12th Python in science conference vol. 13 20 (Citeseer, 2013).

95. Abadi, M., et al. TensorFlow: Large-scale machine learning on heterogeneous systems. (2015).

96. Chollet, F. & Others. keras. (2015).

97. Kotikalapudi, R. & Others. keras-vis. 2017. URL https://github.com/raghakot/keras-vis (2019).

98. Alber, M. et al. iNNvestigate neural networks. J. Mach. Learn. Res. 20, 1–8 (2019).

99. Hossain, S., Calloway, C., Lippa, D., Niederhut, D. & Shupe, D. Visualization of Bioinformatics Data with Dash Bio. in Proceedings of the 18th Python in Science Conference 126–133 (2019).

100. Kohavi, R. & Others. A study of cross-validation and bootstrap for accuracy estimation and model selection. in Ijcai vol. 14 1137–1145 (Montreal, Canada, 1995).

101. Bergstra, J. S., Bardenet, R., Bengio, Y. & Kégl, B. Algorithms for Hyper-Parameter Optimization. in Advances in Neural Information Processing Systems 24 (eds. Shawe-Taylor, J., Zemel, R. S., Bartlett, P. L., Pereira, F. & Weinberger, K. Q.) 2546–2554 (Curran Associates, Inc., 2011).

102. Bergstra, J., Yamins, D. & Cox, D. Making a Science of Model Search: Hyperparameter Optimization in Hundreds of Dimensions for Vision Architectures. in (eds. Dasgupta, S. & McAllester, D.) vol. 28 115–123 (PMLR, 2013).

103. Bergstra, J. & Bengio, Y. Random search for hyper-parameter optimization. J. Mach. Learn. Res. 13, 281–305 (2012).

104. Kingma, D. P. & Ba, J. Adam: A Method for Stochastic Optimization. arXiv [cs.LG] (2014).

105. Zeiler, M. D. ADADELTA: An Adaptive Learning Rate Method. arXiv [cs.LG] (2012).

106. Hinton, G. Slide 29 of Lecture 6, Geoffrey Hinton coursera’s class. http://www.cs.toronto.edu http://www.cs.toronto.edu/~tijmen/csc321/slides/lecture_slides_lec6.pdf.

107. Nair, V. & Hinton, G. E. Rectified Linear Units Improve Restricted Boltzmann Machines. (2010).

108. Klambauer, G., Unterthiner, T., Mayr, A. & Hochreiter, S. Self-Normalizing Neural Networks. in Advances in Neural Information Processing Systems 30 (eds. Guyon, I. et al.) 971–980 (Curran Associates, Inc., 2017).

109. Prechelt, L. Early Stopping - But When? in Neural Networks: Tricks of the Trade (eds. Orr, G. B. & Müller, K.-R.) 55–69 (Springer Berlin Heidelberg, 1998).

110. Hochreiter, S. Untersuchungen zu dynamischen neuronalen Netzen. Diploma, Technische Universität München 91, (1991).

111. Hochreiter, S., Bengio, Y., Frasconi, P., Schmidhuber, J. & Others. Gradient flow in recurrent nets: the difficulty of learning long-term dependencies. (2001).

112. Alqaraawi, A., Schuessler, M., Weiß, P., Costanza, E. & Berthouze, N. Evaluating saliency map explanations for convolutional neural networks: a user study. in Proceedings of the 25th International Conference on Intelligent User Interfaces 275–285 (Association for Computing Machinery, 2020).

113. Selvaraju, R. R. et al. Grad-cam: Visual explanations from deep networks via gradient-based localization. in Proceedings of the IEEE international conference on computer vision 618–626 (2017).

114. Zhou, B., Khosla, A., Lapedriza, A., Oliva, A. & Torralba, A. Learning deep features for discriminative localization. in Proceedings of the IEEE conference on computer vision and pattern recognition 2921–2929 (2016).

115. Wang, Z. & Yang, J. Diabetic Retinopathy Detection via Deep Convolutional Networks for Discriminative Localization and Visual Explanation. arXiv [cs.CV] (2017).

116. Duffy, B. A., et al. Regression activation mapping on the cortical surface using graph convolutional networks. (2019).

117. Le Goallec, A. & Patel, C. J. Age-dependent co-dependency structure of biomarkers in the general population of the United States. Aging 11, 1404–1426 (2019).

