## Supplementary Information for "Comparing the genetic and environmental architecture of blood count, blood biochemistry and urine biochemistry biological ages with machine learning"

### Supplementary Methods, Figures, Tables and References

#### Table of contents

**Abstract** **1**

**Introduction** **2**

**Results** **4**

UKB blood biochemistry biomarkers outperformed urine biochemistry and blood cell biomarkers as age predictors 4

Identification of the laboratory biomarkers driving age prediction 6

Blood biochemistry 6

Blood cells 6

Urine biochemistry 7

Genetic factors and heritability of accelerated aging 7

Blood biochemistry 9

Blood cells 10

Urine biochemistry 10

Biomarkers, clinical phenotypes, diseases, environmental and socioeconomic variables associated with accelerated aging 11

Biomarkers associated with accelerated blood biochemistry aging 12

Clinical phenotypes associated with accelerated blood biochemistry aging 13

Diseases associated with accelerated blood biochemistry aging 14

Environmental variables associated with accelerated blood biochemistry aging 15

Socioeconomic variables associated with accelerated blood biochemistry aging 16

Correlation between blood cell, blood biochemistry and urine biochemistry aging 17

Predicting accelerated aging from biomarkers, clinical phenotypes, diseases, environmental variables and socioeconomic variables 20

**Discussion** **21**

Blood biochemistry outperformed blood cells and urine biochemistry as an age predictor 21

Comparison between our age predictors and the literature 22

Laboratory biomarkers 22

Blood biochemistry 23

Urine biochemistry 24

Blood cells 24
Blood biochemistry, blood cells and urine biochemistry aging are three distinct phenotypes 25
Blood and urine features driving age prediction 25
Blood biochemistry 26
Urine biochemistry 26
Blood cells 26
Accelerated aging is partially heritable 27
Accelerated blood biochemistry aging is linked to aging in other organ systems 29
Environmental and social and economic factors associated with accelerated blood biochemistry aging 29
Limitations 30
**Methods 31**
Data and materials availability 31
Software 31
Cohort Dataset: Participants of the UK Biobank 31
Data types and Preprocessing 32
Machine learning algorithms 32
Training, tuning and predictions 33
Interpretability of the machine learning predictions 33
Ensembling to improve prediction and define aging dimensions 34
Evaluating the performance of models 34
Biological age definition 35
Genome-wide association study of accelerated aging 36
Identification of SNPs associated with accelerated aging 36
Heritability and genetic correlation 37
Non-genetic correlates of accelerated aging 38
**Author Contributions 39**
**Acknowledgments 39**
**Conflicts of Interest 40**
**Funding 40**
**References 41**
**Table of contents 53**
**Methods 56**
Hardware 56
Software 56

Training, tuning and predictions 56
Data splitting 56
Nested cross-validation 58
Bayesian hyperparameters optimization 58
Example 59
Generating average predictions for each participant 61
Interpretability of the predictions 63
Non-genetic correlates of accelerated aging 63
Imputation of the non-genetic X-variables 64
X-Wide Association Studies 65
Prediction of accelerated aging 66
X-Correlations between aging dimensions 66
X-Correlations based on the XWAS results 66
X-Correlations based on the feature importances 67
**Supplementary Figures 68**
**Supplementary Tables 69**
**Supplementary References 75**

### Methods

#### Hardware

We performed the computation for this project on Harvard Medical School's compute cluster, with access to both central processing units [CPUs] and general processing units [GPUs] (Tesla-M40, Tesla-K80, Tesla-V100) via a Simple Linux Utility for Resource Management [SLURM] scheduler.

#### Software

We coded the project in Python [1] and used the following libraries: NumPy [2,3], Pandas [4], Matplotlib [5], Plotly [6], Python Imaging Library [7], SciPy [8–10], Scikit-learn [11], LightGBM [12], XGBoost [13], Hyperopt [14], TensorFlow 2 [15], Keras [16], Keras-vis [17], iNNvestigate [18]. We used Dash [19] to code the website on which we shared the results. We set the seed for the os library, the numpy library, the random library and the tensorflow library to zero.

#### Training, tuning and predictions

##### Data splitting

We split the 676,787 samples into ten data folds, while keeping all samples from the same participant in the same fold. To ensure this, we split the 502,211 participants' ids (referred to by UKB as "eid") into ten different buckets of the same size. To generate ten folds for each sub-dataset (e.g. urine biochemistry), we took the intersection of the samples in each of the ten folds with the samples for which the sub-dataset data was available. This method had however one important loophole, which is that we could not guarantee that the folds for the sub-datasets would be balanced. For example, urine microalbumin levels were only measured for 156,589 out of the 502,211 participants. Since the 502,211 participants are split into ten folds, a fold contains approximately 50,221 participants. Although unlikely, we could therefore not guarantee that all or most of the urine microalbumin samples would be attributed to the first four data folds, leading to highly unbalanced folds for the urine biochemistry analysis. Unbalanced folds can lead to problems during the cross-validation (see further below), as models trained on a smaller number of samples will tend to generalize worse. One solution would have been to use a different split for each dataset, but this would have generated problems when building the ensemble models fold by fold (see Methods - Models ensembling). To mitigate this issue of unbalanced data folds, we developed the following heuristic. We randomly split the 502,211 participants into ten folds, 1,000 times. For each of these 1,000 splits, we computed for each sub-dataset the variance of the percentages of samples in each fold. We then scored each of the 1,000 splits using the maximum of the variance among the different sub-datasets. For example, if the urine biochemistry samples were not evenly split for the ith split out of the 1,000 splits (e.g. fold 1: 55% of the samples, every other fold: 5% of the samples), the variance of the sample proportions would be high, which would yield a poor score for the ith split. Finally, we selected the split with the lowest score as the final split for the main dataset, and for all the sub-datasets. This selected split had a score of 5.8e-4, which means that the most unbalanced sub-dataset had a variance in its sample size proportion between its ten folds of 5.8e-4.

#### Nested cross-validation

Cross-validation is a method to tune the regularization of models and prevent overfitting [20]. For the models inputting scalar data (Figure 1A in green), we tuned the hyperparameters and generated a testing prediction for each sample using a nested 10x9-folds cross-validation. We refer to the two nested cross-validations as the “outer” and the “inner” cross-validations. The outer-cross validation is used to generate an unbiased testing prediction for each sample, as opposed to a simple split of the data into a “training+validation” set on one hand, and a testing set on the other hand, which would only generate a testing prediction for one tenth of the dataset. The inner cross-validation is used to tune the hyperparameters more precisely, leveraging the full inner cross-validation dataset as a validation set, as opposed to a simple data split of the “training+validation” dataset into a training and a validation sets, which would only use one data fold as the validation set to estimate the performance associated with a specific combination of hyperparameters. The nested cross-validation is illustrated in Table S27.

#### Bayesian hyperparameters optimization

To tune the hyperparameters, we used the Tree-structured Parzen Estimator Approach [21] [TPE] of the hyperopt python package [22]. TPE is a sequential Bayesian hyperparameters optimization method that iteratively suggests the next most promising hyperparameters combination as a function of the hyperparameters combinations that have already been tested, by building a probabilistic representation of the objective function. We set the number of iterations to 30. For each model, 30 different hyperparameter combinations are iteratively tested before selecting the best performing one. The hyperparameters names and their ranges defining the hyperparameters space can be found in Table S26. It might be of interest to other researchers that we initially tuned the hyperparameters using a random search [23] with the same number of iterations, and we did not observe a significant improvement in the model's performance after implementing the Bayesian hyperparameters optimization.

#### Example

For the sake of clarity, let us walk through a concrete example, which is illustrated in Table S27. Suppose we want to generate unbiased predictions for every sample in a dataset using an elastic net. First, let us generate the testing prediction for the data fold F9, which is performed by the first fold of the outer cross-validation (outer cross-validation fold 0). We select the data fold F9 out of the ten data folds as the testing fold, and we select the remaining nine data folds as "training+validation" folds for the inner cross-validation. We scale and center the target (age) and the predictors using the mean and standard deviation values of the variables on the "training+validation" dataset. We then enter the first inner-cross validation.

For the first inner cross-validation fold, we select the data fold F8 as the validation set, and the remaining eight "training+validation" data folds as the training set. We re-scale and center age and the predictors in the training and the validation sets using the mean and standard deviation values of the training set. We train the model on the eight training data folds with the first hyperparameters combination sampled by the TPE algorithm (one value for alpha and one value for l1_ratio) and generate validation predictions on the validation fold (data fold F8), which we unscale. This completes the first of the nine inner cross-validation folds (Inner CV fold 0). We then permute the nine inner data folds. We scale the age and the predictors using the mean and standard deviation computed on the new training set. Then we train the model with the same first combination of hyperparameters on eight data folds, leaving aside the data fold F9 (still being used as the testing set for the outer cross-validation) and the data fold F7 (now being used as the validation set for the inner cross-validation). We then use the new trained model to generate validation predictions on the data fold F7, which we unscale. This completes the second of the nine inner-cross validation folds (Inner CV fold 1). We then reiterate these inner permutation and training processes seven more times, until every data fold in the nine “training+validation” data folds is used as the validation set once. At this point, we concatenate the validation predictions from these nine validation folds to obtain the overall validation predictions associated with the first hyperparameters combination, and compute the associated performance metric (e.g. RMSE). This completes the inner-cross validation for the first hyperparameters combination.

We then perform the same 9-folds inner cross-validation, this time with the second hyperparameters combination suggested by the TPE algorithm. We iterate this process 28 more times, until 30 different hyperparameters combinations have iteratively been tested. Next, we select the hyperparameter combination that yielded the best validation performance (e.g. minimum RMSE), and we retrain a model on the whole nine “training+validation” data folds (all data folds except for data fold #1), using this best performing hyperparameters combination. This completes the first inner cross-validation.

We then use the model to generate unbiased predictions on the unseen testing set (data fold F9) and record these predictions. By anticipation for the ensembling algorithm (see Methods - Models ensembling) we also need to compute validation predictions on the data fold F8. We do this by training a model on all the data folds aside from the validation fold (data fold F8) and the testing fold (data fold F9), with the selected hyperparameters combination. We then use this trained model to compute predictions on the validation fold (data fold F8) and record these predictions, after unscaling them. This completes the first of the ten outer cross-validation folds (outer cross-validation 0).

We then complete the second outer cross-validation fold (outer cross-validation 1), this time using the data fold F8 as the testing dataset, to obtain unbiased testing predictions on this data fold, as well as validation predictions on the data fold F7. We reiterate the process eight more times to obtain the testing and validation predictions on the remaining data folds. We then concatenate the testing predictions from the ten data folds to obtain our final testing predictions for the model. Similarly, we concatenate the validation predictions from the ten data folds to obtain our final testing predictions for the model, which will later be used during ensemble models building and model selection (see Methods - Models ensembling).

The final validation and testing predictions for each data fold are therefore not necessarily associated with the same hyperparameters combination. It is also important to notice that we performed a single outer cross-validation, but that we performed a separate inner-cross validation for each outer cross-validation fold (hence the word “nested”), for a total of ten inner cross-validations per outer cross-validation fold.

#### Generating average predictions for each participant

We generated an average prediction for each individual, reported to UKB’s instance 0. We walk through an example. Let us assume a participant had two glucose levels samples collected from them in instances 2 and 3, respectively at age 70 and 80. Let us assume that the age predictions were respectively 64 and 78, so the residuals are respectively -6 years and -2 years, for an average of -4 years. However, we still need to take into account the bias in the residuals, defined as the difference between the participant's chronological age and the prediction. As explained in more details under Methods - Biological age definition, we observed a bias in the residuals as a function of chronological age. Participants on the younger end of the chronological age distribution tend to be predicted older than they actually are, whereas participants on the older end of the distribution tend to be predicted younger than they actually are. We need to properly account for this bias when translating a prediction from a more recent instance to an older instance. Let us assume that the average bias in the residuals for participants who are 70 and 80 years old is respectively -2 years and -4 years. After correcting for this bias, the predictions are now respectively 64-(-2)=66 and 78-(-4) = 82. Therefore, the corrected residuals for this participant are respectively -4 years and +2 years, for an average of -1 years. Finally, let us assume that the participant was 60 years old in instance 0. We will assign a single prediction of 60-1=59 years to the participant, but we still need to un-correct for the bias in residuals. Let us assume that the average bias for the residuals at age 60 is +5 years. We will assign a final prediction for the participant of 59+5=64 years.

A key point we would like to highlight here is that we did not actually correct for the bias in the residuals at this step of the pipeline. Instead, we corrected then un-corrected the predictions that we translated from different instances to the instance 0. The actual correction for the residual biases takes place when defining the biological age phenotypes (see Methods - Biological age definition).

To distinguish between raw predictions on the instance 0, and the average predictions reported to the instance 0, we created a new instance which we named instance "*". We refer to these predictions as "participants predictions", as opposed to "samples predictions".

#### Interpretability of the predictions

For elastic nets, we interpreted the models using the values of the regression coefficients. Large absolute values for these coefficients means they played an important role when generating the predictions. For gradient boosted machines we used the feature importances, which are based on the number of times a tree selected each of the variables. Variables with high feature importances were selected more often and are therefore likely to play a key role in predicting chronological age. For neural networks, we estimated the importance of each feature by permuting it randomly between samples before computing the performance of the model. The score of each feature is the difference between the R-Squared value before and after the random permutations. Features whose random permutation leads to a large decrease in the model's performance are estimated to be important predictors of chronological age.

We estimated the concordance between the three different algorithms by computing the Pearson and the Spearman correlations between their feature importances.

#### Non-genetic correlates of accelerated aging

Unlike DNA, biomarkers, phenotypes, diseases, family history, environmental variables and socioeconomics can change over life. As a consequence, we compared each biomarker, phenotype and environmental variable with the accelerated aging of the participant at the time the exposure was measured and we used the "Samples predictions", as opposed to the "Participants predictions" that we used for the identification of genetic correlates (see Methods - Models ensembling - Generating average predictions for each participant).

#### Imputation of the non-genetic X-variables

Most X-variables were not collected on all four instances. Additionally, no X-variables were collected at the same time as the accelerometer data was collected. To identify the non-genetic correlates of accelerated aging, we had to impute the values of the X-variables for the ages of the participants for which they were not available. We considered two imputation methods, which we refer to as the “cross-sectional” and the “longitudinal” imputations.

For the cross-sectional imputation, we computed a linear regression for each X variable as a function of age, adjusting for sex. We then used the slope of the linear regression to extrapolate the value of the XWAS variable at different ages.

For the longitudinal imputation, we first selected, for each X variable, all the participants that had at least two measures taken for this X variable. We then performed a linear regression for each participant. We then averaged the slope of the linear regressions over all the participants of the same sex. Finally, we used this slope to extrapolate the value of the XWAS variable at different ages for all participants depending on their sex, in the same way we did it for the cross-sectional imputation.

It is important to notice that for both the cross-sectional imputation and the longitudinal imputation, data can only be imputed when the XWAS variable has been measured at least once for the participant. This raw measure is then used to extrapolate which value the X variable was likely taking a couple years earlier and/or later.

The advantage of the cross-sectional imputation is larger sample sizes. The advantage of the longitudinal method is that it corrects for generational effects. For example, old people have shorter legs than young people on average [24]. This is not because human legs shrink as we grow older. Instead, people who are old today already had shorter legs when they were young. If the cross-sectional regression is used to impute the length of the participants on instances where it was not measured, it will spuriously assign smaller values to the older samples. In contrast, the longitudinal regression learns the regression coefficient by comparing each participant to themselves as they age and will therefore not capture the generational effect. When used to predict the participants legs' length, it will impute constant values over time. To evaluate which of the two imputation methods should be preferred, we used them to predict X-variables for which we knew the actual values and computed the R-Squared values associated with the predictions. We found that, even with sample sizes as small as 200 samples, longitudinal imputation outperformed cross-sectional imputation. We therefore used longitudinal imputation.

#### X-Wide Association Studies

First, we tested for associations in an univariate context by computing the partial correlation between each X-variable and aging dimensions. To compute the partial correlation between an X-variable and an aging, we followed a three steps process. (1) We ran a linear regression on each of the two variables, using age, sex and ethnicity as predictors. (2) We computed the residuals for the two variables. (3) We computed the correlation between the two residuals and the associated p-value if their intersection had a sample size of at least ten samples. We used a threshold for significance of 0.05 and corrected the p-values for multiple testing using the

Bonferroni correction. We plotted the results using a volcano plot. We refer to this pipeline as an X-Wide Association study [XWAS].

In the supplementary tables and the results, we rank the X-variables subcategories by decreasing percentage of variables associated with accelerated aging (note that the ranking is therefore biased towards categories with fewer variables). For each subcategory, we list the three most associated variables, based on the absolute value of the correlation coefficient. For the exhaustive list, please refer to https://www.multidimensionality-of-aging.net/xwas/univariate_associations.

#### Prediction of accelerated aging

We leveraged the pipeline we built to predict chronological age as a function of scalar biomarkers to predict accelerated aging for the different aging dimensions as a function of the biomarkers, clinical phenotypes, diseases, family history, environmental and socioeconomic variables. We leveraged the same pipeline to identify which features were driving the predictions. We built a model for each X-variables subcategory (Table S7, Table S10, Table S13, Table S16, Table S19, Table S22).

#### X-Correlations between aging dimensions

##### X-Correlations based on the XWAS results

For the sake of clarity, let us walk through an example. We want to compute the environmental correlation between accelerated blood biochemistry and blood cells aging. The XWAS generates a vector whose components are the partial correlations between the accelerated aging phenotype and each environmental variable, for both accelerated blood biochemistry and blood cells aging. We compute three different Pearson correlations between these two partial correlation vectors. (1) The “All” correlation, using all the components of the two vectors; this correlation tends to be inflated by the large number of X-variables whose correlation with both accelerated aging dimensions is close to zero. (2) The “Intersection” correlation, using only the environmental variables that were significantly associated with both of the accelerated aging dimensions;because the cardinality of the intersection can be small, a small number of X-variables can yield very high or very low correlations. (3) The “Union” correlation, using only the environmental variables that were significantly associated with at least one of the two accelerated aging dimensions; the “Union” correlation represents a compromise between the “All” and the “Intersection” correlations. The figures in this paper were generated using the “Union” correlation, but all three correlations can be explored on the website.

##### X-Correlations based on the feature importances

We then computed the correlations between the feature importances for different accelerated aging dimensions to estimate the X-correlation between the different dimensions. We used the same method as described above under “X-Correlations based on the XWAS results”, replacing the coefficient obtained for each X-variable in a univariate context (using partial correlation with accelerated aging) with the coefficient obtained in a multivariate context (as an accelerated aging predictor in a multivariate model).

### Supplementary Figures

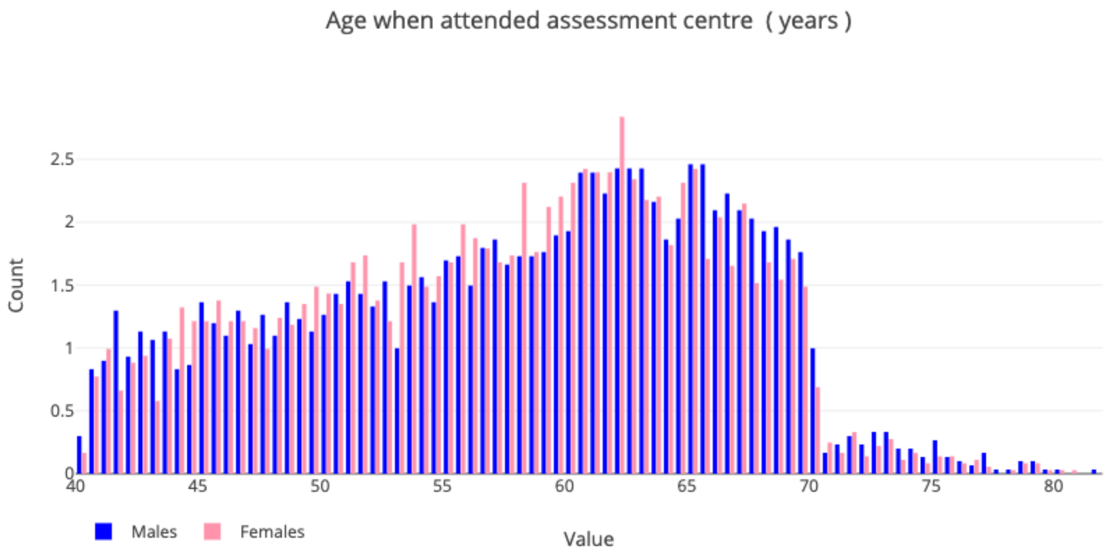


**Figure S1: Demographics of the UK Biobank cohort**

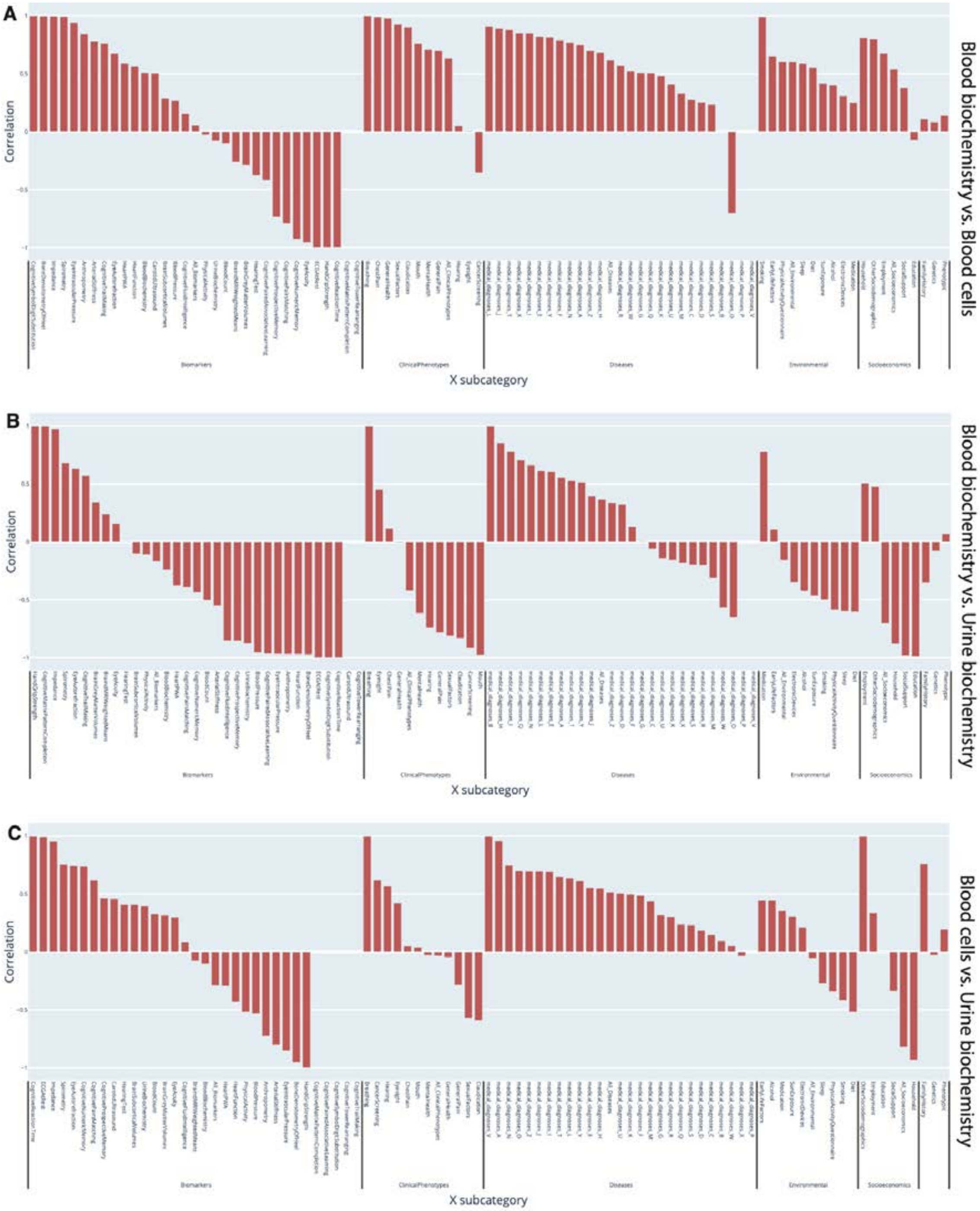


**Figure S2: Correlation between accelerated aging in the three aging dimensions in terms of phenotype, genetics, biomarkers, clinical phenotypes, disease, environment and socioeconomics. A - Blood biochemistry vs. Blood cells. B - Blood biochemistry vs. Urine Biochemistry. C - Blood cells vs. Urine biochemistry.**

### Supplementary Tables

**Table S1: Comparison between the performance of the models**

| Aging dimension | Number of Predictors (non-demographics) | Elastic Net (R-Squared) | GBM (R-Squared) | Neural Network (R-Squared) |
|---|---|---|---|---|
| Blood biochemistry | 28 | 0.302±0.005 | 0.476±0.005 | 0.474±0.004 |
| Urine biochemistry | 4 | 0.081±0.003 | 0.104±0.003 | 0.096±0.003 |
| Blood cells | 31 | 0.058±0.004 | 0.122±0.002 | 0.119±0.003 |

**Table S2: Feature importances for the models built on blood biochemistry biomarkers**

See supplementary data

**Table S3: Feature importances for the models built on blood cells biomarkers**

See supplementary data

**Table S4: Feature importances for the models built on urine biochemistry biomarkers**

See supplementary data

**Table S5: Pearson correlations between the feature importances for different scalar features-based algorithms**

| Aging dimension | Correlation vs. ElasticNet | Correlation vs. GBM | Correlation vs. NeuralNetwork | ElasticNet vs. GBM | ElasticNet vs. NeuralNetwork | GBM vs. NeuralNetwork |
|---|---|---|---|---|---|---|
| Blood biochemistry | 0.595 | 0.406 | 0.143 | 0.601 | 0.185 | -0.102 |
| Urine biochemistry | 0.721 | 0.508 | 0.491 | 0.821 | 0.257 | 0.073 |
| Blood cells | 0.322 | 0.228 | 0.407 | 0.43 | 0.075 | -0.099 |

**Table S6: Spearman correlations between the feature importances for different scalar features-based algorithms**

| Aging dimension | Correlation vs. ElasticNet | Correlation vs. GBM | Correlation vs. NeuralNetwork | ElasticNet vs. GBM | ElasticNet vs. NeuralNetwork | GBM vs. NeuralNetwork |
|---|---|---|---|---|---|---|
| Blood biochemistry | 0.491 | 0.412 | 0.344 | 0.792 | 0.294 | 0.316 |
| Urine biochemistry | 0.411 | 0.446 | 0.595 | 0.697 | 0.379 | 0.441 |
| Blood cells | 0.354 | 0.271 | 0.585 | 0.394 | 0.241 | 0.228 |

**Table S7**: **List of biomarkers by subcategories for the Biomarkers Wide Association Study [BWAS]**

See supplementary data

**Table S8: Biomarkers most associated with accelerated aging for each aging dimension**

See supplementary data

**Table S9: Biomarkers most associated with decelerated aging for each aging dimension**

See supplementary data

**Table S10: List of clinical phenotypes by subcategories for the Clinical Phenotypes Wide Association Study [CWAS]**

See supplementary data

**Table S11: Clinical phenotypes most associated with accelerated aging for each aging dimension**

See supplementary data

**Table S12: Clinical phenotypes most associated with decelerated aging for each aging dimension**

See supplementary data

**Table S13: List of diseases by subcategories for the Diseases Wide Association Study [DWAS]**

See supplementary data

**Table S14: Diseases most associated with accelerated aging for each aging dimension**

See supplementary data

**Table S15: Diseases most associated with decelerated aging for each aging dimension**

See supplementary data

**Table S16: List of family history variables by subcategories for the Family History Phenotypes Wide Association Study [FWAS]**

See supplementary data

**Table S17: Family history variables most associated with accelerated aging for each aging dimension**

See supplementary data

**Table S18: Family history variables most associated with decelerated aging for each aging dimension**

See supplementary data

**Table S19: List of environmental variables by subcategories for the Environmental Wide Association Study [EWAS]**

See supplementary data

**Table S20: Environmental variables most associated with accelerated aging for each aging dimension**

See supplementary data

**Table S21: Environmental variables most associated with decelerated aging for each aging dimension**

See supplementary data

**Table S22: List of socioeconomic variables by subcategories for the Socioeconomics Wide Association Study [SWAS]**

See supplementary data

**Table S23: Socioeconomic variables most associated with accelerated aging for each aging dimension**

See supplementary data

**Table S24: Socioeconomic variables most associated with decelerated aging for each aging dimension**

See supplementary data

**Table S25: Correlation between accelerated aging in the three aging dimensions in terms of phenotype, genetics, biomarkers, clinical phenotypes, disease, environment and socioeconomics**

| Dimension 1 | Dimension 2 | Phenotypic | Genetic | Biomarkers | Clinical phenotypes | Diseases | Environment | Socioeconomics |
|---|---|---|---|---|---|---|---|---|
| Blood biochemistry | Blood cells | .143+-.002 | .085+-0.011 | .056 | .636 | .618 | .603 | .539 |
| Urine biochemistry | Blood biochemistry | .070+-.003 | -.080+-.043 | -.170 | .423 | .369 | -.160 | -.704 |
| Blood cells | Urine biochemistry | .195+-.003 | -.031+-.025 | -.291 | -.035 | .516 | -.057 | -.824 |

**Table S26: Hyperparameter space for scalar features-based models Bayesian optimization**

| Algorithm | Hyperparameter | Scale | Low | High |
|---|---|---|---|---|
| Elastic net | alpha | loguniform | -10 | 0 |
| | l1_ratio | uniform | 0 | 1 |
| Gradient Boosted Machine | num_leaves | quniform | 5 | 45 |
| | min_child_samples | quniform | 100 | 500 |
| | min_child_weight | loguniform | -5 | 4 |
| | subsample | uniform | 0.2 | 0.8 |
| | colsample | uniform | 0.4 | 0.6 |
| | reg_alpha | loguniform | -2 | 2 |
| | reg_lambda | loguniform | -2 | 2 |
| | n_estimators | quniform | 150 | 450 |
| Neural network | learning_rate_init | loguniform | -5 | -1 |
| | apha | loguniform | -6 | 3 |

### Table S27: Nested Cross-Validation pipeline

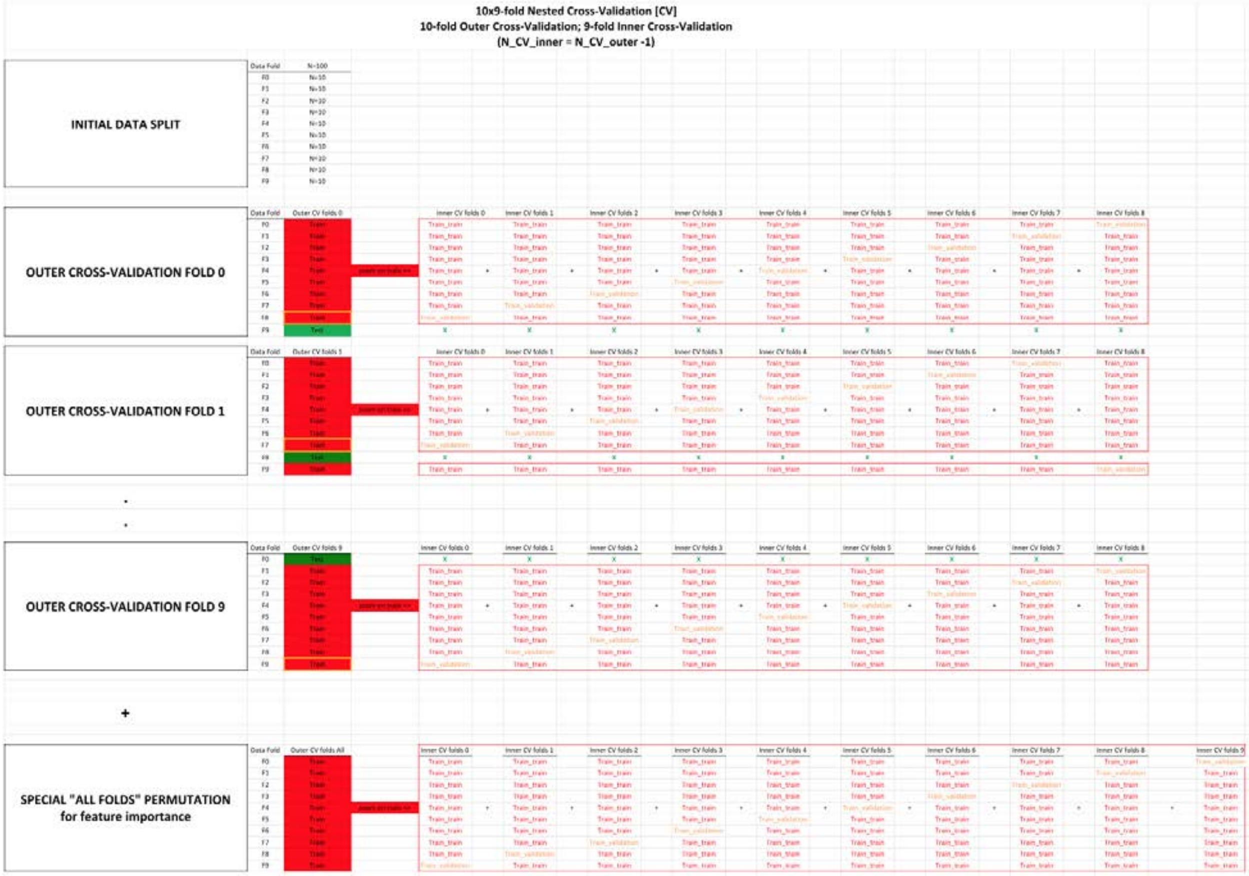


| METHODS: | |
|---|---|
| General comments | The Nested Cross-Validation is a normal double split train/val/test method, for which not just one, but the two splits are replaced with a CV instead. |
| Outer Cross-Validation | The Outer Cross-Validation is used to replace the split between train + val on one side, vs. test on the other side. Thanks to the Outer Cross-Validation, an unbiased testing prediction can be obtained for every sample.<br>Three common sources of confusion clarified:<br>**Yes,** using an Outer Cross-Validation does mean that the predictions do not all come from the same model. (For exemple for testing, the prediction for each fold comes from a different model.)<br>**No**, the Outer Cross-Validation has ***nothing*** to do with hyperparameter tuning or model selection.<br>Tuning a model only requires one Outer Cross-Validation. In contrast, it requires N_CV_folds distinct Inner Cross-Validations, one for each fold of the Outer Cross Validation. (see below) |
| Inner Cross-Validation | The Inner Cross-Validation is used to replace the split between the typical training set and validation set.<br>Thanks to the Inner Cross-Validation, hyperparameter selection can be decided based on a performance calculated on the full training set and validation set sample size, instead of the sole validation set sample size.<br>Two common sources of confusion clarified:<br>#1 - The whole purpose of performing an Inner Cross-Validation is ***hyperparameter selection***.<br>The Inner Cross-Validation tunes the model that will then be used to generate predictions on the left out testing fold.<br>Therefore, the Inner Cross-Validation does ***not*** generate a testing prediction for more samples. That is the role of the Outer Cross-Validation.<br>One last time, the Inner Cross-Validation is ***only*** about improving the hyperparameter tuning of the model.<br>#2 - Tuning a model requires N_CV_folds distinct Inner Cross-Validations, one for each fold of the Outer Cross Validation. In contrast, it only requires one Outer Cross-Validation. (see above) |
| Special Fold 0 | Special "All folds" permutation is only here for feature importance purposes.<br>We could extract the features importance from the 10 models used to generate the predictions on the testing set, but we would have to take the average of features importances between several model.<br>Additionally each of these models only saw 90% of the data (10% was left aside for testing purposes).<br>If we only care about feature importance, we actually do not need to leave samples aside for testing. We can use 100% of the data to train a model using a 10-fold (not 9-fold this time, since the test fold can be used too!) ***Inner*** Cross-Validation to train a single model.<br>This model will never be tested, but it does not matter: its role is to provide the best estimate for the feature importance, which it does well.<br>Two common sources of confusion clarified:<br>#1 - The Nested Cross-Validation pipeline and the Special "All Folds" permutation pipeline are ***not*** intertwined and serve different purposes.<br>The purpose of the Nested Cross-Validation pipeline is to generate accurate and a unbiased testing prediction for every sample, whereas the purpose of the Special "All Folds" pipeline is to generate feature importance.<br>#2 - **Yes**, it does mean that the feature importance reported do not correspond to any of the 10 models (trained in the 10 Outer Cross-Validation folds) that generated the final testing predictions.<br>The feature importance comes from a model that was never used to generate predictions. |

| RESULTS: | **Example**: cells corresponding to the values used for each prediction set | **Explanation**: |
|---|---|---|
| Training | "E17+G17+I17+K17+M17+O17+Q17+S17+ E29+G29+I29+K29+M29+O29+Q29+U29+..." | To obtain the training prediction for the Data Fold F1, take the mean of every training prediction available on this data fold (10*8=80 of them). |
| Validation | "E25" | To obtain the validation prediction for the Data Fold F1, take the prediction outputted by the model trained on the remaining 8 data folds with the best hyperparameters values obtained for this Outer Cross-Validation fold.<br>These validation predictions can then be used to tune the ensemble models. /!\ Warning /!\: This can only be properly done if data folds split used for the Outer Cross-Validation and the Inner Cross-Validation is the same. |
| Testing | "C26" | No extra step is needed for the testing prediction for the Data Fold 1. Simply take the only testing prediction available for this fold. |
| Feature importance: | N/A | Extract the features importance from the special "outer fold" "All".<br>If want to compute the standard deviation for the feature importance as well, we can leverage the feature importance computed on each of the Outer Cross-Validation folds and take the square root of their variance. |

**SUPPLEMENTARY:**

**Comparison between Nested Cross-Validation and other validation methods:**

**#1**-Single split train/test.

Split train/test works when there are no hyperparameters to tune. If one uses a simple train/test split and tune the hyperparameters on the testing set, some overfitting is to be expected.

**#2**-Double split train/validation/test.

Because of the above, a train/validation/test double split is commonly used when hyperparameters tuning is required.

The training is performed on the training set, the tuning of the hyperparameters on the validation set, and the evaluation of the unbiased model performanc on the untouched test dataset.

**#3**-Cross-Validation(train/val) and split (train+val)/test.

With the pipeline #2 (double split train/validation/test), the tuning of the hyperparameters is not leveraging the dataset as much as with a Cross-Validation, as only one data fold is used to tune the hyperparameters.

Therefore machine learning practitioners usually replace the "inner split" (the training/validation split), by a Cross-Validation instead, but they keep the "outer split" (between the training+validation dataset used for their Cross-Validation and the testing set).

**#4**-Split train/val and Cross-Validation((train+val)/test).

Pipeline #3 only generate an unbiased testing prediction for a fraction of the dataset (the testing set).

To generate an unbiased prediction on every sample of the dataset, pipeline #4 can be used.

However, because there is a simple split between the training set and the validation set (one for each Cross-Validation fold), that means that the hypperparameter tuning only leverages a fraction of the dataset.

Pipeline #4 is preferred when generating an unbiased testing prediction for every sample is needed but the models are too time consuming to train to be able to afford a Nested Cross-Validation (#5).

Pipeline #4 is therefore the pipeline we used to predict chronological age using medical images or videos, for example.

**#5**-Nested Cross-Validation.

To fully leverage the whole dataset, a Nested Cross-Validation replaces both the outer split (train+val/test) and the inner splits (train/val) by a Cross-Validation, hence the nesting.

It requires the training of a large number of models, and it is therefore usually reserved for models that can be trained relatively quickly.

In our case, we used it to train our scalar features-based models (elastic nets, light gradient boosted machines, and shallow fully connected neural networks).

### Supplementary References


1. Van Rossum, G. & Drake, F. L. *The Python Language Reference Manual*. (Network Theory Limited, 2011).
2. Oliphant, T. E. *A guide to NumPy*. vol. 1 (Trelgol Publishing USA, 2006).
3. Walt, S. van der, van der Walt, S., Chris Colbert, S. & Varoquaux, G. The NumPy Array: A Structure for Efficient Numerical Computation. *Computing in Science & Engineering* vol. 13 22–30 (2011).
4. McKinney, W. & Others. Data structures for statistical computing in python. in *Proceedings of the 9th Python in Science Conference* vol. 445 51–56 (Austin, TX, 2010).
5. Hunter, J. D. Matplotlib: A 2D Graphics Environment. *Comput. Sci. Eng.* **9**, 90–95 (2007).
6. Inc, P. T. Collaborative data science. *Montreal: Plotly Technologies Inc Montral* (2015).
7. Clark, A. Pillow Python Imaging Library. *Pillow—Pillow (PIL Fork) 5. 4. 1 documentation* (2018).
8. Virtanen, P. *et al.* SciPy 1.0: fundamental algorithms for scientific computing in Python. *Nature Methods* vol. 17 261–272 (2020).
9. Oliphant, T. E. Python for Scientific Computing. *Computing in Science Engineering* **9**, 10–20 (2007).
10. Millman, K. J., Jarrod Millman, K. & Aivazis, M. Python for Scientists and Engineers. *Computing in Science & Engineering* vol. 13 9–12 (2011).
11. Pedregosa, F. *et al.* Scikit-learn: Machine learning in Python. *the Journal of machine Learning research* **12**, 2825–2830 (2011).
12. Ke, G. *et al.* LightGBM: A Highly Efficient Gradient Boosting Decision Tree. in *Advances in*

*Neural Information Processing Systems 30* (eds. Guyon, I. et al.) 3146–3154 (Curran Associates, Inc., 2017).

13. Chen, T. & Guestrin, C. XGBoost: A Scalable Tree Boosting System. in *Proceedings of the 22nd ACM SIGKDD International Conference on Knowledge Discovery and Data Mining* 785–794 (Association for Computing Machinery, 2016).
14. Bergstra, J., Yamins, D. & Cox, D. D. Hyperopt: A python library for optimizing the hyperparameters of machine learning algorithms. in *Proceedings of the 12th Python in science conference* vol. 13 20 (Citeseer, 2013).
15. Abadi, M. *et al.* TensorFlow: Large-scale machine learning on heterogeneous systems. (2015).
16. Chollet, F. & Others. keras. (2015).
17. Kotikalapudi, R. & Others. keras-vis. 2017. *URL https://github. com/raghakot/keras-vis* (2019).
18. Alber, M. *et al.* iNNvestigate neural networks. *J. Mach. Learn. Res.* **20**, 1–8 (2019).
19. Hossain, S., Calloway, C., Lippa, D., Niederhut, D. & Shupe, D. Visualization of Bioinformatics Data with Dash Bio. in *Proceedings of the 18th Python in Science Conference* 126–133 (2019).
20. Kohavi, R. & Others. A study of cross-validation and bootstrap for accuracy estimation and model selection. in *Ijcai* vol. 14 1137–1145 (Montreal, Canada, 1995).
21. Bergstra, J. S., Bardenet, R., Bengio, Y. & Kégl, B. Algorithms for Hyper-Parameter Optimization. in *Advances in Neural Information Processing Systems 24* (eds. Shawe-Taylor, J., Zemel, R. S., Bartlett, P. L., Pereira, F. & Weinberger, K. Q.) 2546–2554 (Curran Associates, Inc., 2011).
22. Bergstra, J., Yamins, D. & Cox, D. Making a Science of Model Search: Hyperparameter

Optimization in Hundreds of Dimensions for Vision Architectures. in (eds. Dasgupta, S. & McAllester, D.) vol. 28 115–123 (PMLR, 2013).

23. Bergstra, J. & Bengio, Y. Random search for hyper-parameter optimization. *J. Mach. Learn. Res.* **13**, 281–305 (2012).

24. Le Goallec, A. & Patel, C. J. Age-dependent co-dependency structure of biomarkers in the general population of the United States. *Aging* **11**, 1404–1426 (2019).
